# Childhood body composition and BMI as a predictor of cardiometabolic non-communicable diseases in adulthood: A systematic review

**DOI:** 10.1101/2021.05.22.21251399

**Authors:** Amela Bander, Alexia J Murphy-Alford, Victor O Owino, Cornelia U Loechl, Jonathan CK Wells, Imara Gluning, Marko Kerac

## Abstract

There is growing evidence that childhood malnutrition is associated with non-communicable diseases (NCDs) in adulthood and that body composition mediates some of this association. This review aims to determine: if childhood body composition can be used to predict later-life cardiometabolic NCDs; which measures of body composition best predict future NCDs. Three electronic databases were searched for studies where: children aged under 5 year had body composition measured; cardiometabolic health outcomes were measured a minimum of 10 years later. 29 studies met the inclusion criteria. Though a poor proxy measure of body composition, Body mass index (BMI) was commonly reported (*n*=28, 97%). 25% of these studies included an additional measure (Ponderal Index or skinfold thickness). Only some studies adjusted for current body size (*n*=11, 39%). Many studies reported that low infant BMI and high childhood BMI increased the risk of NCD-related outcomes in later life but no conclusions can be made about exact timing of child malnutrition and consequent impact on NCD. Because studies focused on BMI rather than direct measures of body composition, nothing can be said about which measures of body composition in childhood are most useful. Future research on child nutrition and long-term outcomes is urgently needed and should include validated body composition assessments as well as standard anthropometric and BMI measurements.

## 1. Introduction

Non-communicable diseases (NCDs), such as cardiovascular diseases, diabetes and chronic respiratory diseases, are the leading cause of mortality, equivalent to 71% of deaths worldwide, and are projected to increase even further, reaching 52 million deaths by 2030 [1].

Risk factors for NCDs include both social factors (poverty, education and stress) and biological (e.g. fetal epigenetic changes with life-course consequence): the former highly affects lifestyle factors such as diet and physical activity [2]. Early life malnutrition is also a key risk factor for NCDs and refers to insufficient energy-and/or nutrient intake; but also refers to an excessive and imbalanced energy intake, often resulting in overweight or obesity [3]. For assessing nutritional status in children and adults, anthropometric indicators of growth and body size such as weight-for-height (WHZ), weight-for-age (WAZ), body mass index (BMI) and mid-upper-arm circumference (MUAC) amongst others are commonly used [4]. However, there is growing evidence that anthropometry alone has marked limitations in describing nutrition-related risk (of morbidity/mortality)[5]. Body composition measures – of which there are several – are attracting interest as potentially much better indicators of both short- [6] and long-term risk [7,8].

There is extensive evidence that: exposure to in-utero undernutrition increases the risk of NCDs in later life [9– 11] and being overweight in adulthood increases the risk of NCDs [12,13]. There is also emerging evidence relating to childhood exposures[14], one recent review finding that *“exposure to severe malnutrition or famine in childhood was consistently associated with increased risk of cardiovascular disease, hypertension, impaired glucose metabolism and metabolic syndrome in later life”* [15]. In attempts to better understand the link between such episodes of early-life malnutrition to later life health and NCD, an increasing number of studies are assessing body composition in childhood [16,17]. Whilst plausible [18], the links between body composition in early life and later-life NCDs are not currently well understood [19–24]. Our review thus aims to synthesize evidence on early life body composition and long-term cardiometabolic health, and to examine which measures of body composition best predict the risk of NCDs.

## 2. Materials and Methods

The PRISMA (Preferred Reporting Items for Systematic review and Meta-Analysis) protocol was used for this systematic review [25].

### 2.1 Inclusion/exclusion criteria

Inclusion criteria was based on PICOS outline:

- **Population:** Subjects who had nutritional status (BMI or body composition) measured at baseline at any time from birth up to 5 years of age with a follow-up time ≥10y.
- **Intervention/exposure:** Exposure to any of the following body composition measurements: skinfolds, bioelectrical impedance analysis, dual-energy X-ray absorptiometry (DXA/DEXA) scan, isotope dilution and PEA POD air displacement plethysmography. Despite only being proxy indicators of body composition, we also included body mass index (BMI) and ponderal index (PI).
- **Comparator/control:** Studies with and without a control group are included.
- **Outcome:** Cardiometabolic non-communicable diseases (coronary artery disease, type 2 diabetes, metabolic syndrome) and their associated risk factors (obesity, blood pressure, blood glucose levels, lipid levels, waist circumference) measured ≥10y after exposure.
- **Study design:** All study designs were considered eligible.

The review excluded studies with a high-risk study population, grey literature, unpublished studies, reviews, non-human studies and studies not published in English, in full format and before 1990.

### 2.2 Search strategy

The search was completed independently by two authors in three databases: *Embase Classic + Embase, Ovid MEDLINE (R) and In-Process & Other non-Indexed Citations and Daily*, and *Global Health*. The final search was conducted 27.07.2020. A detailed search strategy is shown in Appendix A.

### 2.3 Study selection

All records generated from the search were imported into Mendeley Reference Manager (version 1.19.4) and were screened by title and abstract. Articles that were deemed relevant or where more information was needed to determine relevance, were screened by full-text.

### 2.4 Data extraction

A data extraction form developed for this review was used to extract information from eligible studies. When obtainable, the following information was extracted: author, year, title, country, study design, sample size, percentage female, inclusion- and exclusion criteria, type of exposure and assessment method, type of outcome and assessment method, years of follow-up, adjustment for current body size, key findings and strength of evidence.

### 2.5 Data analysis

Due to heterogeneity amongst studies identified, the analysis is presented as a narrative synthesis. Results from high-income countries (HIC) and low- and middle-income countries (LMIC) are analysed separately and should not be compared.

### 2.6 Assessing risk of bias

An individual risk of bias assessment for each study was determined using the ‘Quality appraisal checklist for quantitative studies reporting correlation and associations’ in ‘Methods for the development of NICE public health guidance’ [26].

### 2.7 Study protocol

A pre-registered protocol for this review can be found at: https://www.crd.york.ac.uk/prospero/display_record.php?RecordID=188393

## 3. Results

### 3.1 Study selection

Selection process and search results are presented in Figure 1. The search generated 5772 records. Following deduplication and initial screening of titles and abstract, 78 articles were eligible for full-text review. Of these, 49 did not meet the inclusion criteria which led to a total of 29 studies included in the review.

**Figure 1.**
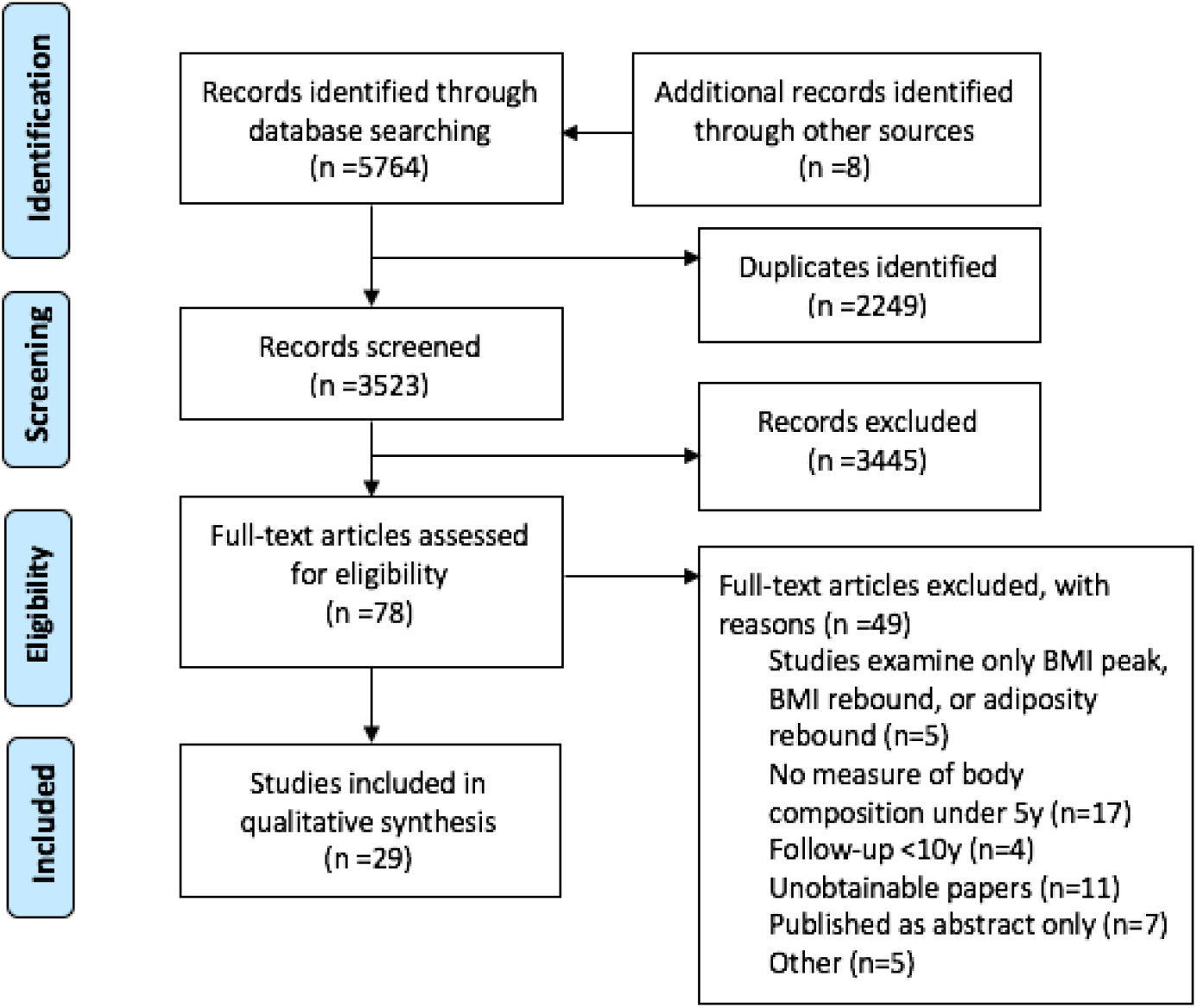
Study selection flow diagram.

### 3.2 Study characteristics

Study characteristics are presented in Table 1. Most studies were from HIC (n=21, 72%), and all but one study used BMI as the indicator of early life exposure / body composition (n=28, 97%). Few studies (n= 7, 25%) used an additional indictor, which were either ponderal index (n=4, 40%) or skinfold thickness (n=3, 38%). No studies used direct measures of body composition: bioelectrical impedance analysis, dual-energy X-ray absorptiometry (DXA/DEXA) scan, isotope dilution and PEA POD air displacement plethysmography).

**Table 1.**
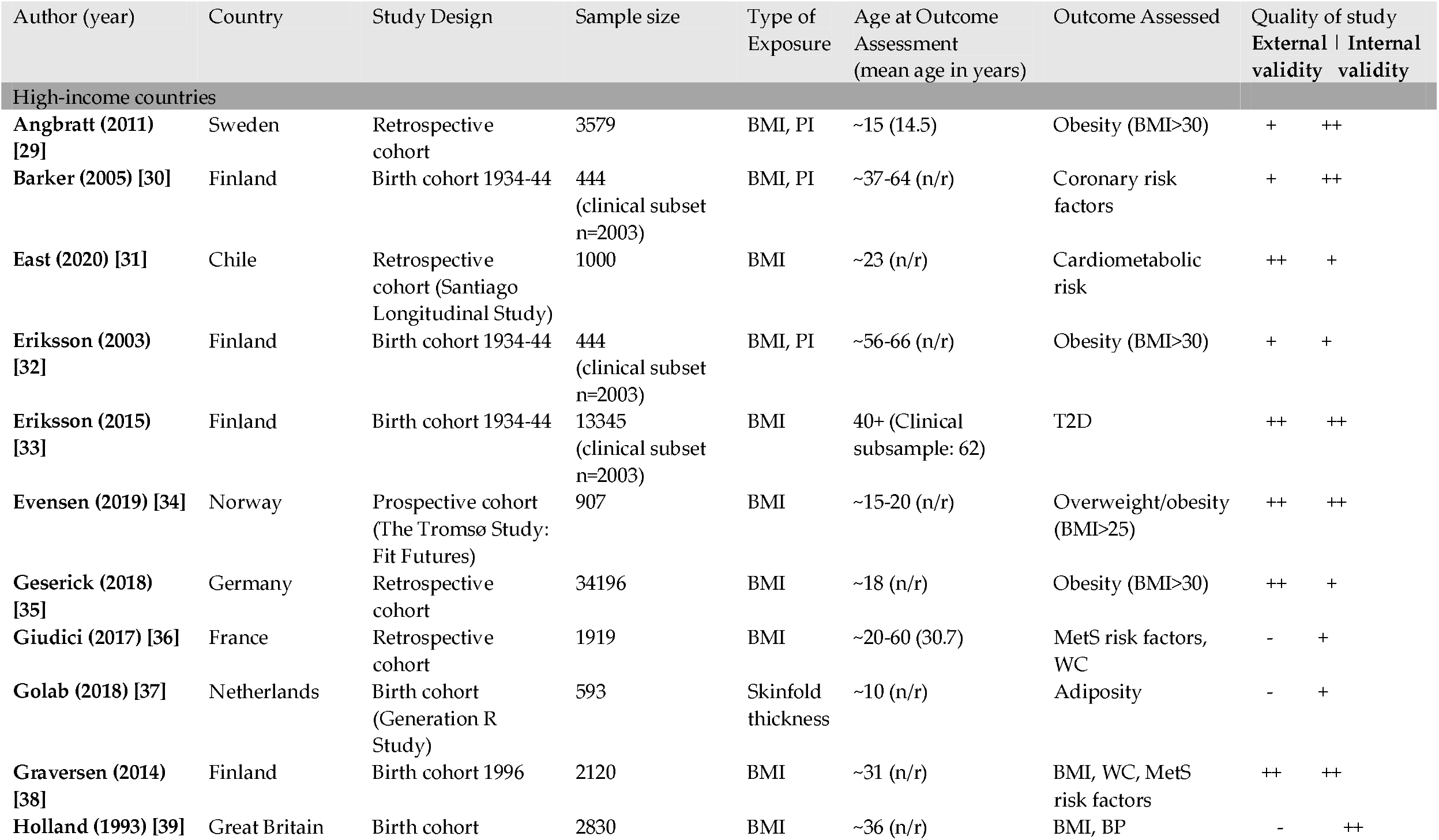
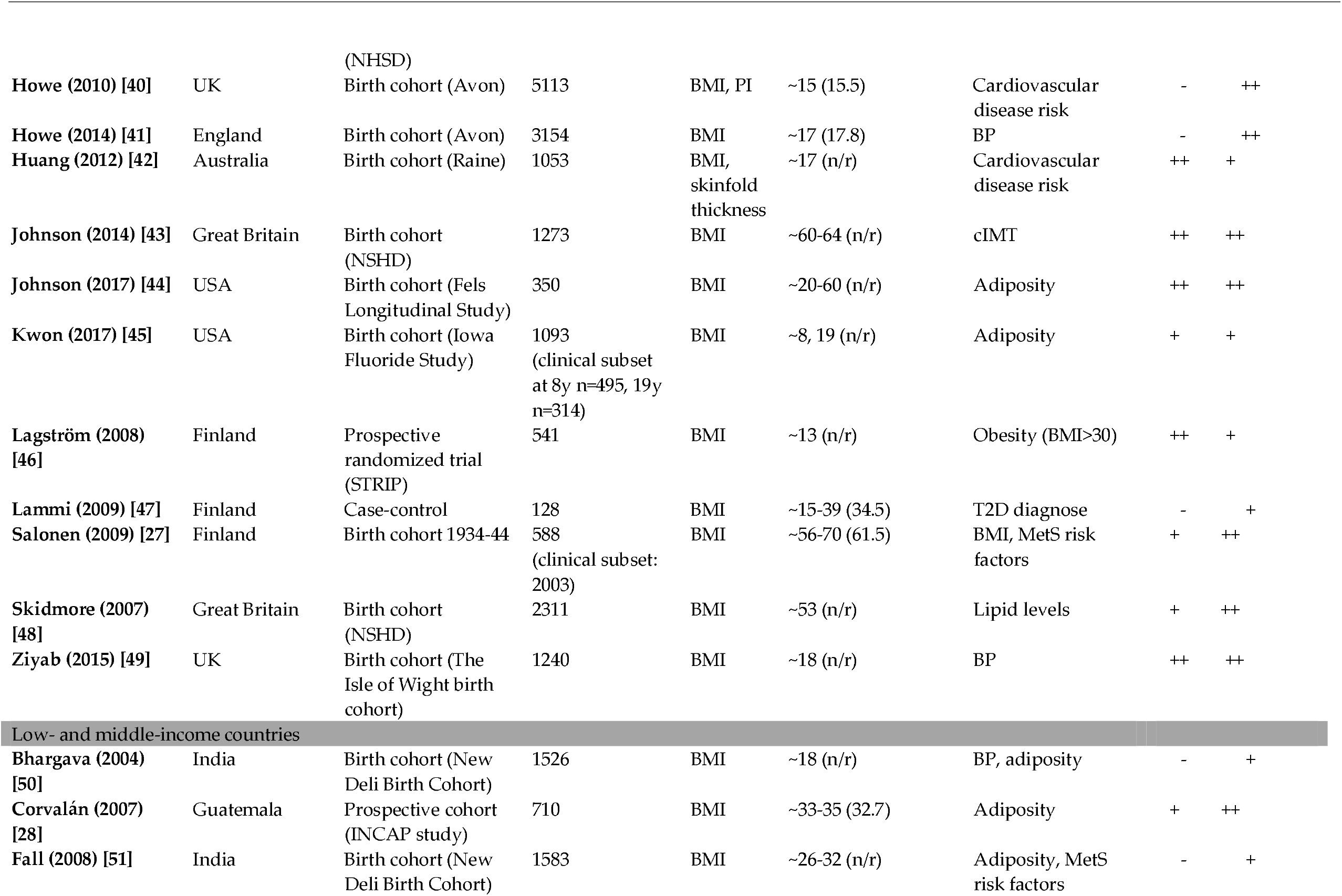
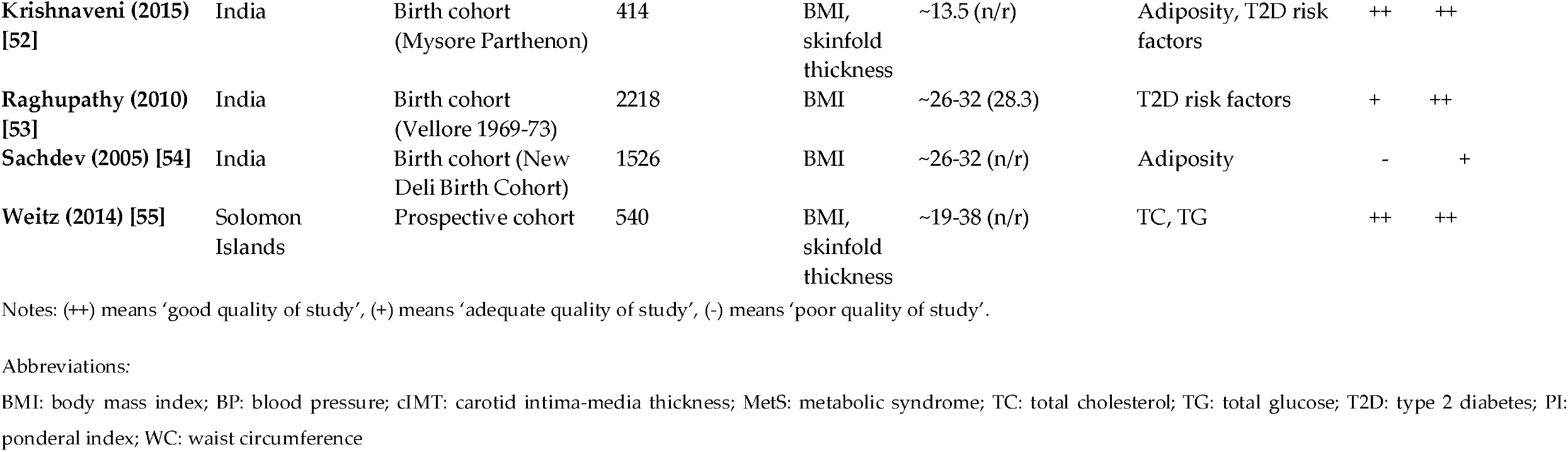
Study characteristics of included stories.

Sample size included in the analysis ranged from 128 to 34196 participants. Participants were most often drawn from existing cohorts (of which 19 were birth cohorts), and 4 studies from HIC recruited the study population from health care registers from the respective countries. All studies were representative cohorts, although the study population in one Finnish study were exclusively normal-weight in adulthood [27] and the Guatemalan study population had a high prevalence of stunting (53% stunted by age 7) [28].

Studies included in the review were a mix of good, adequate and poor quality. External validity for 9 studies was rated to be of poor quality due to various reasons e.g. reduced power and significant proven differences between study population and participants who were lost to follow up.

### 3.3 Synthesis of results

Table 2 presents a summary of included studies. A detailed summary can be found in Appendix B. The following section describes the results of the included studies.

**Table 2.**
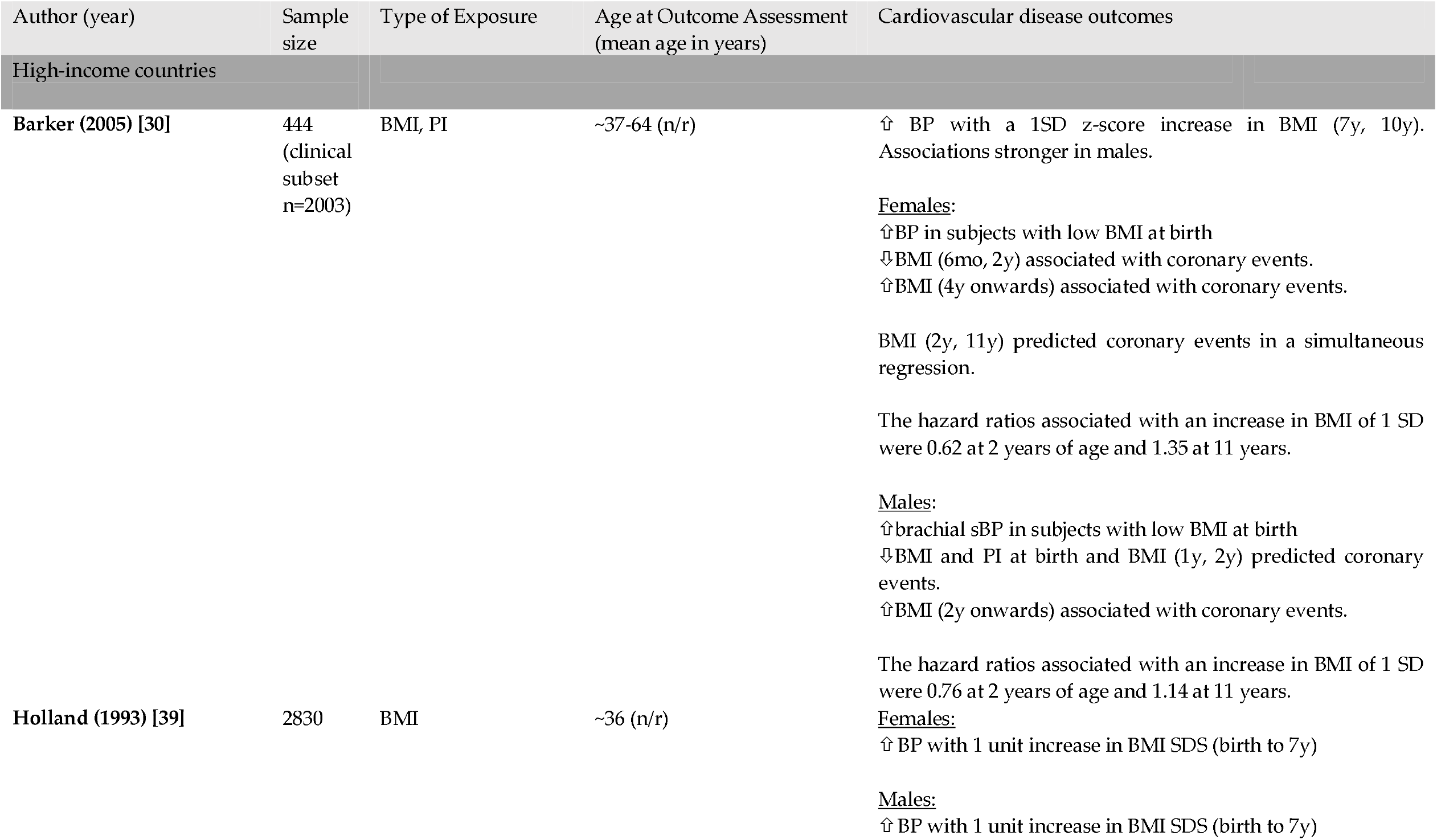
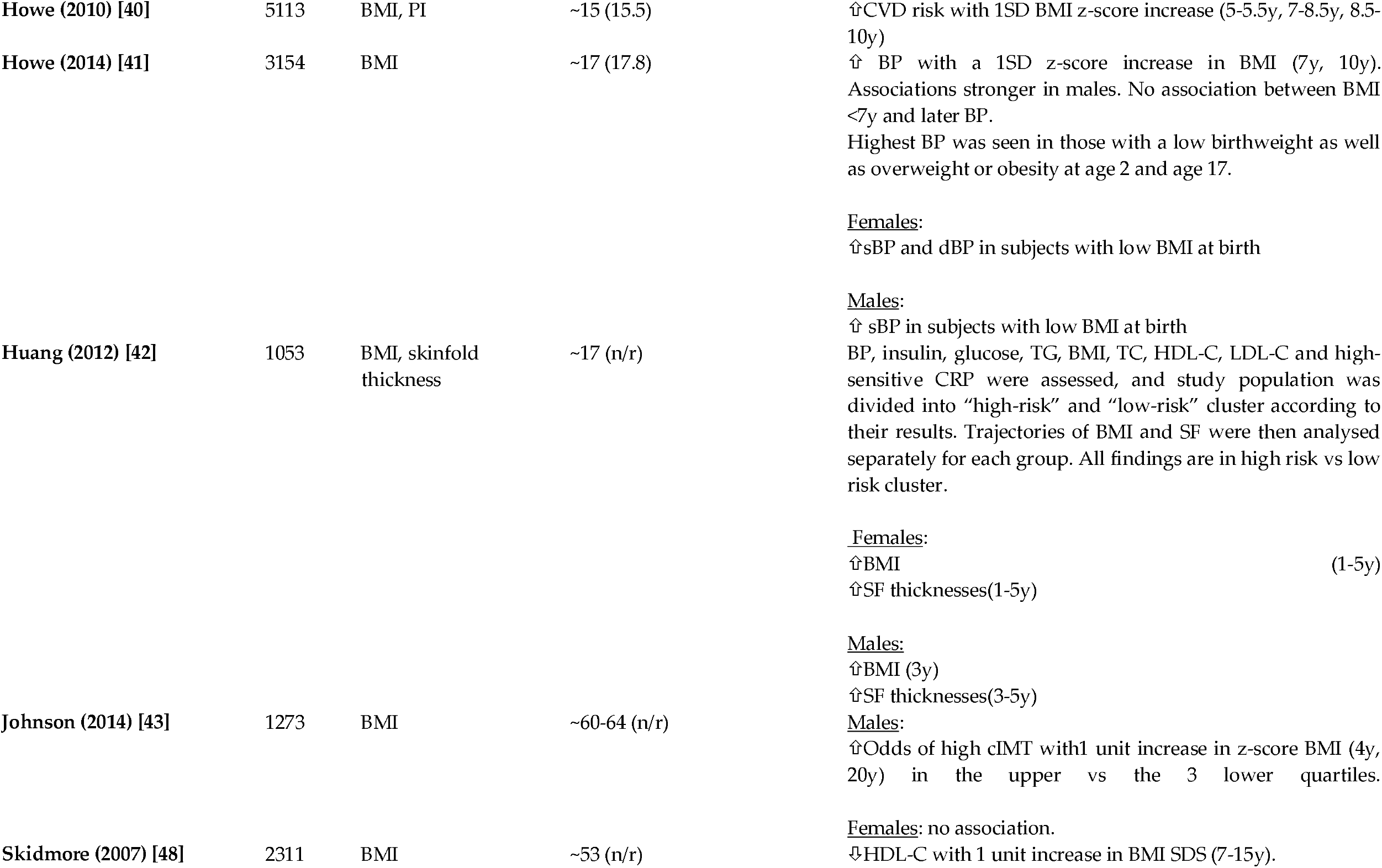
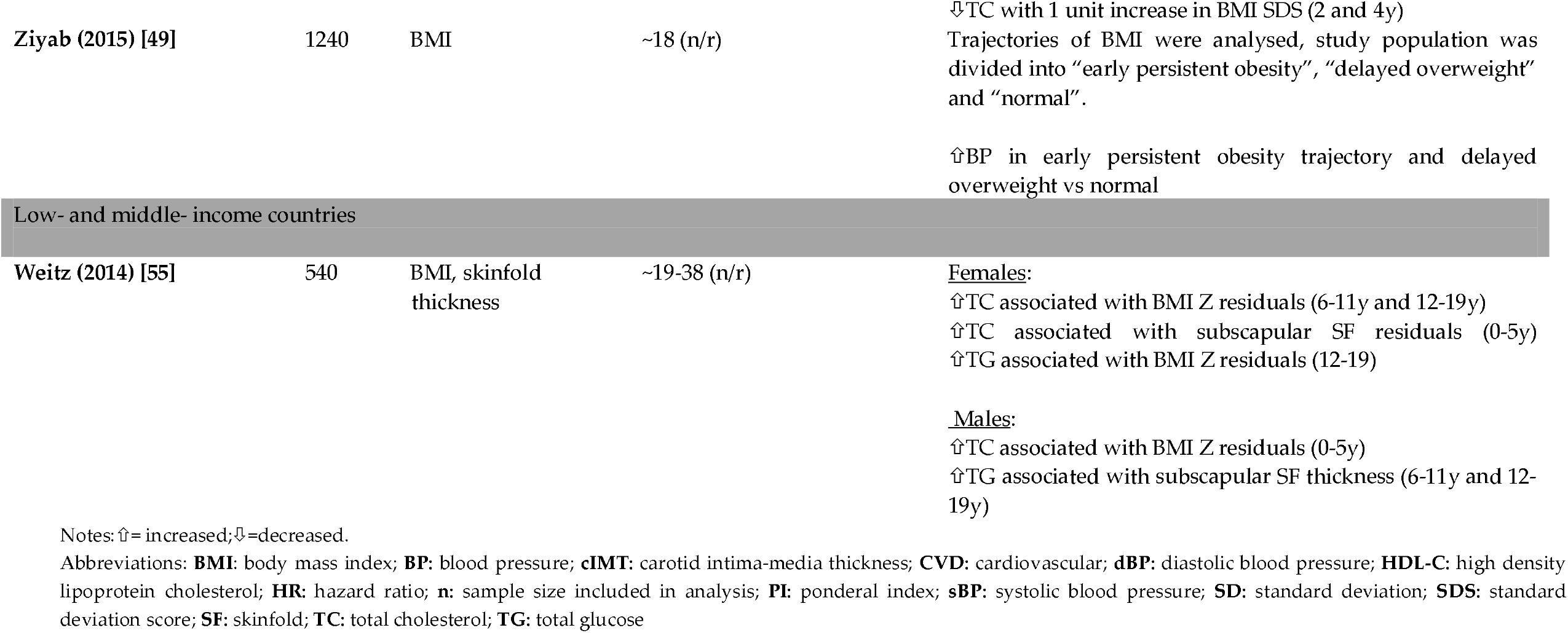
Summary of studies reporting on cardiovascular disease outcomes.

**Table 3.**
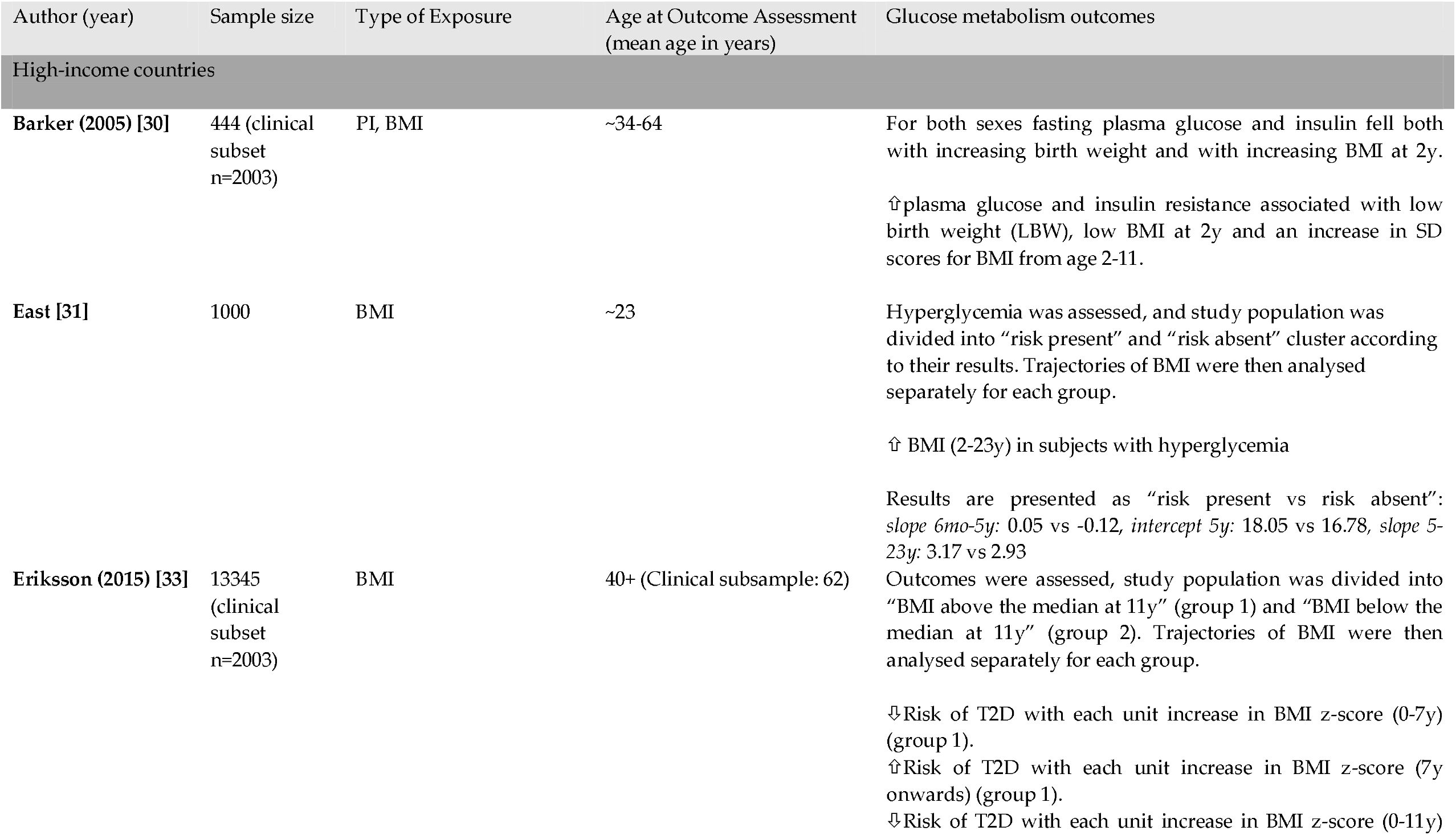
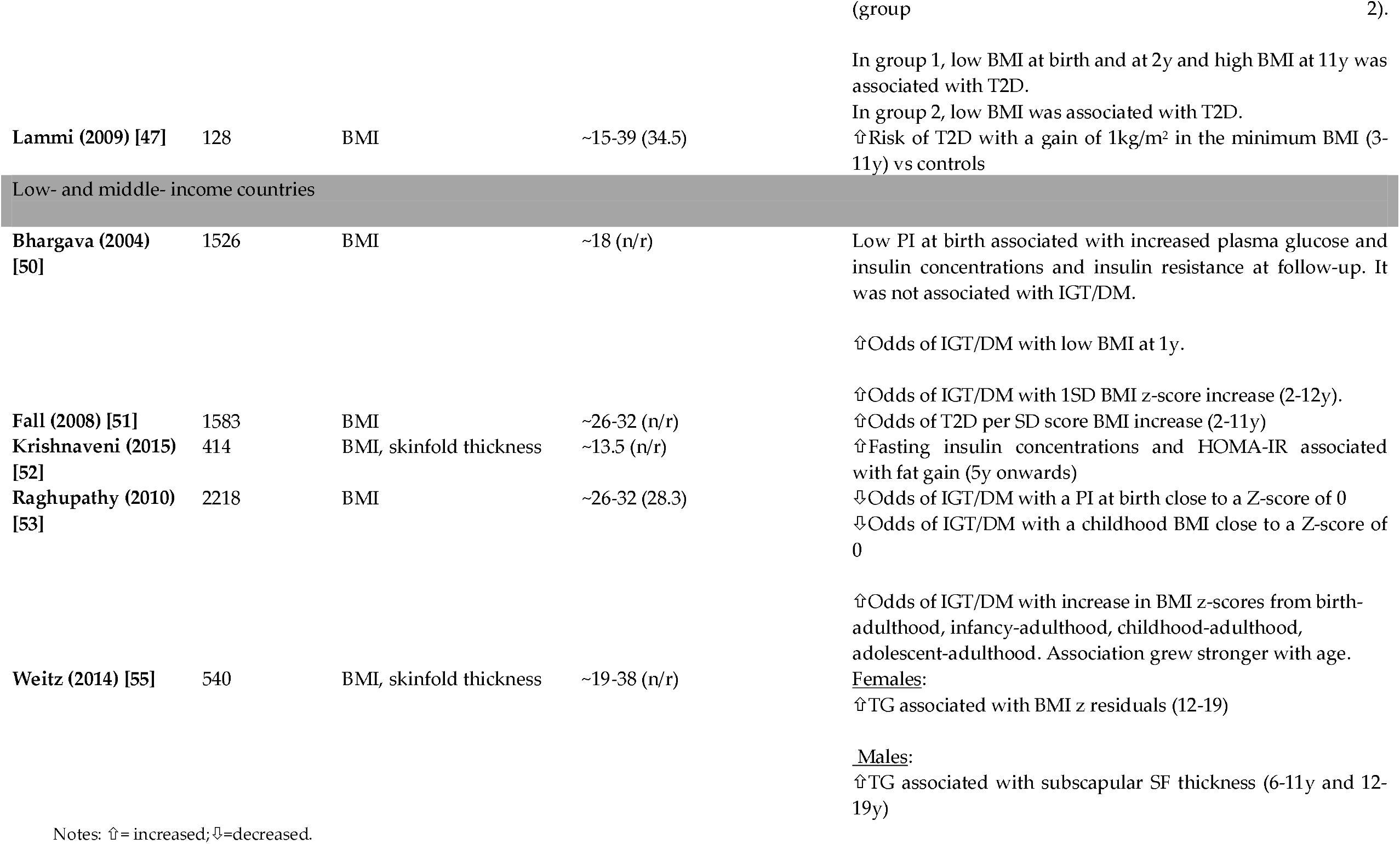
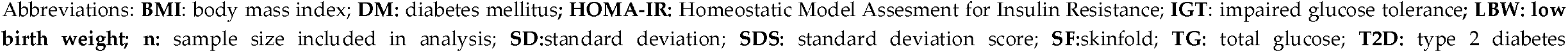
Summary of studies reporting on glucose metabolism outcomes.

**Table 4.**
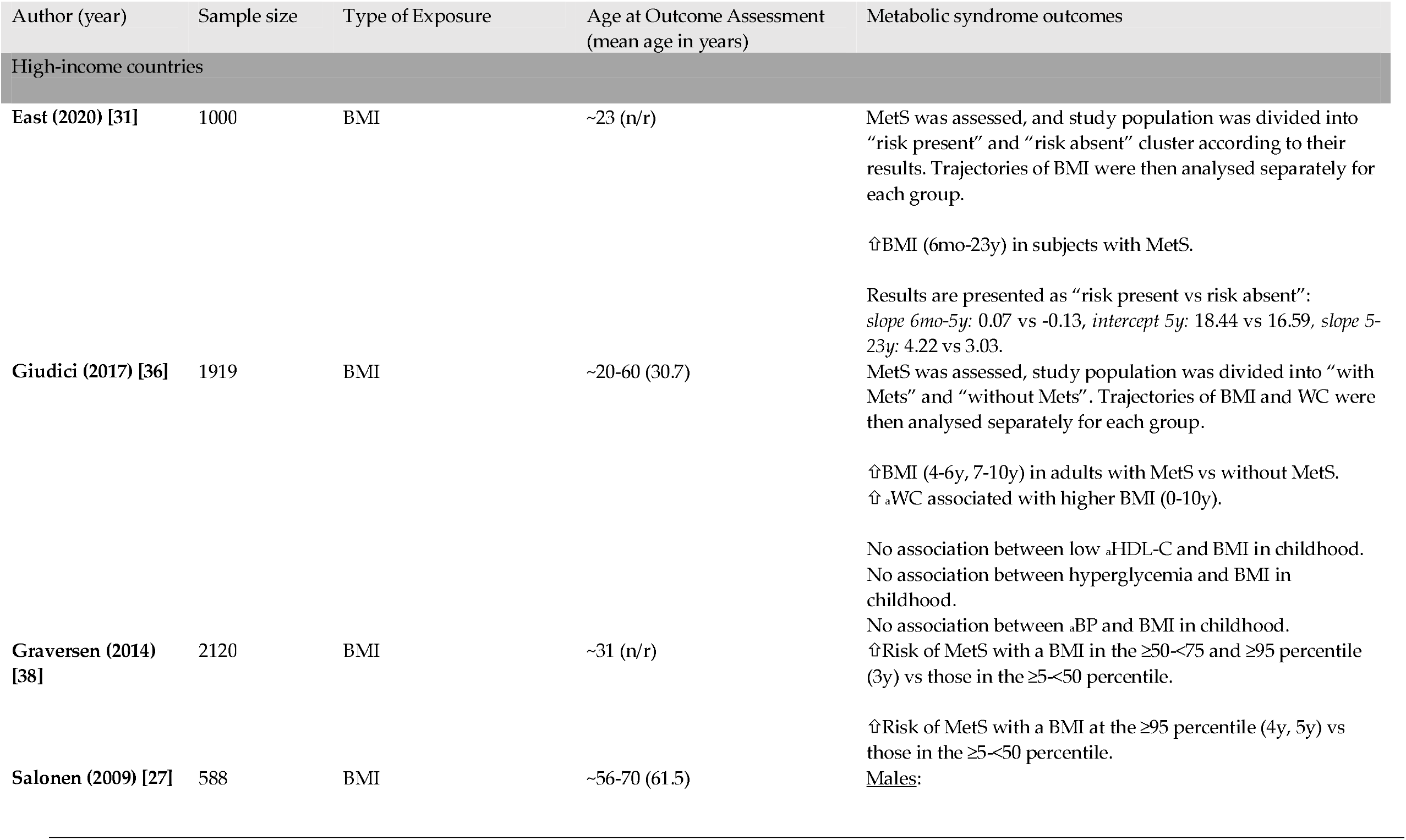
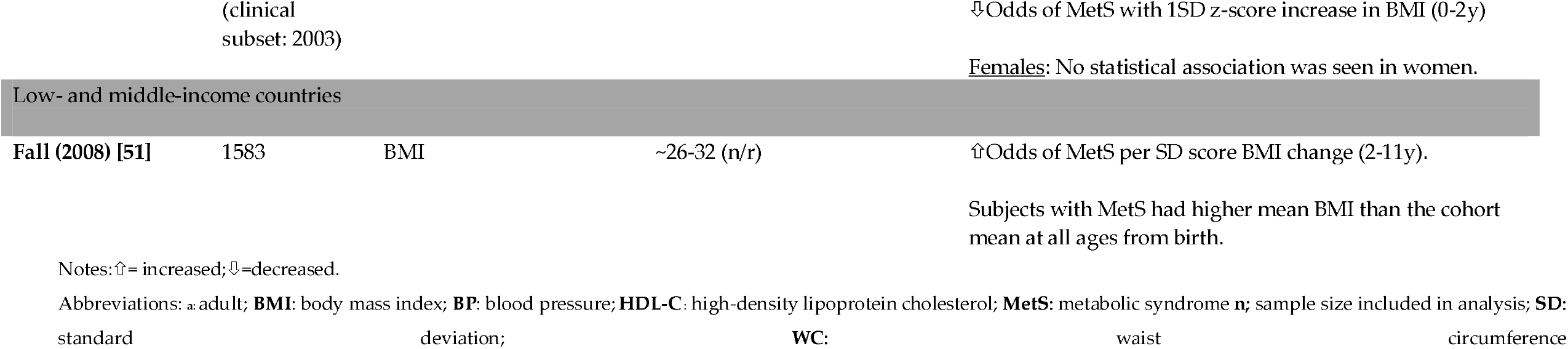
Summary of studies reporting on metabolic syndrome outcomes.

**Table 5.**
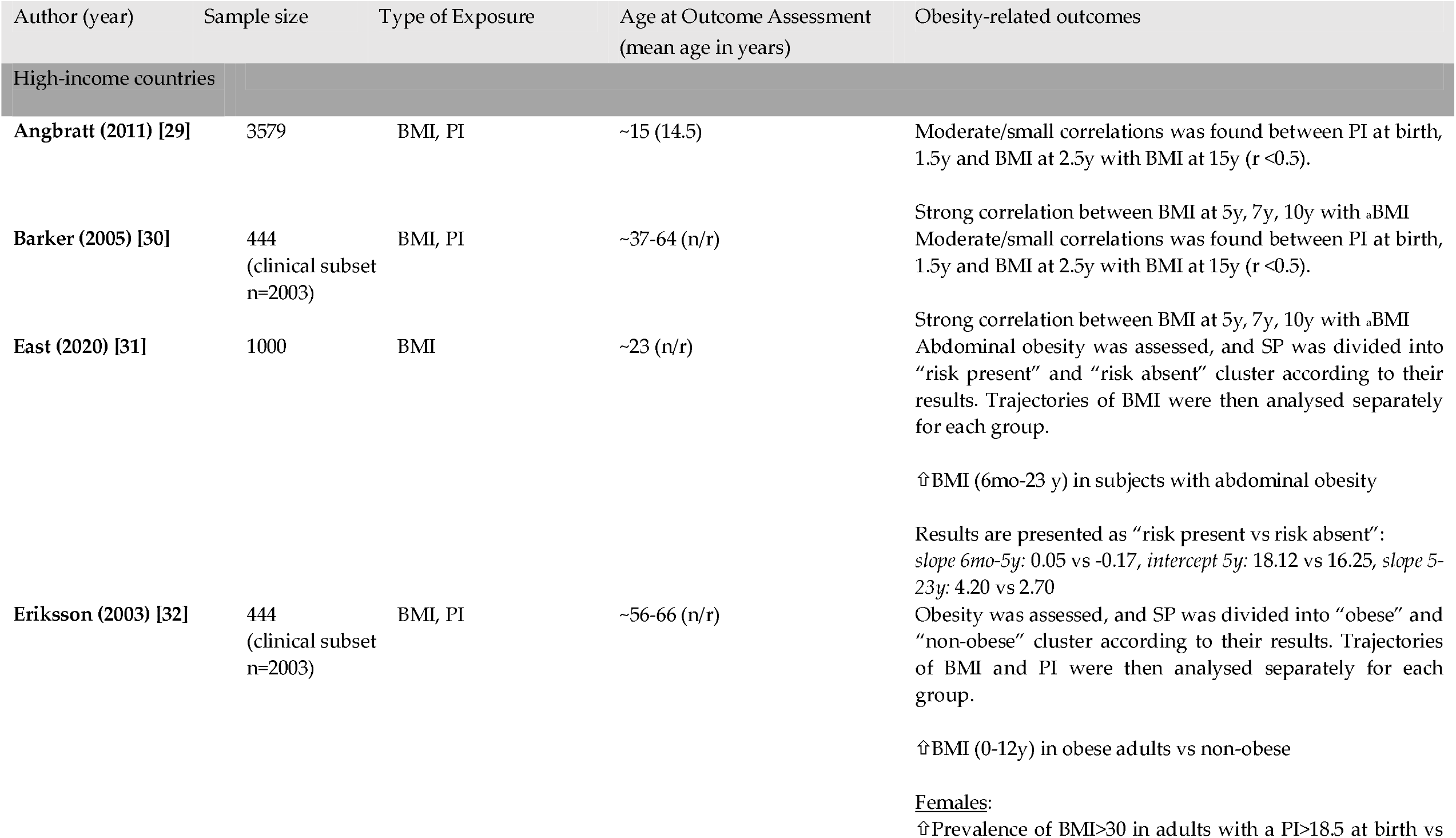
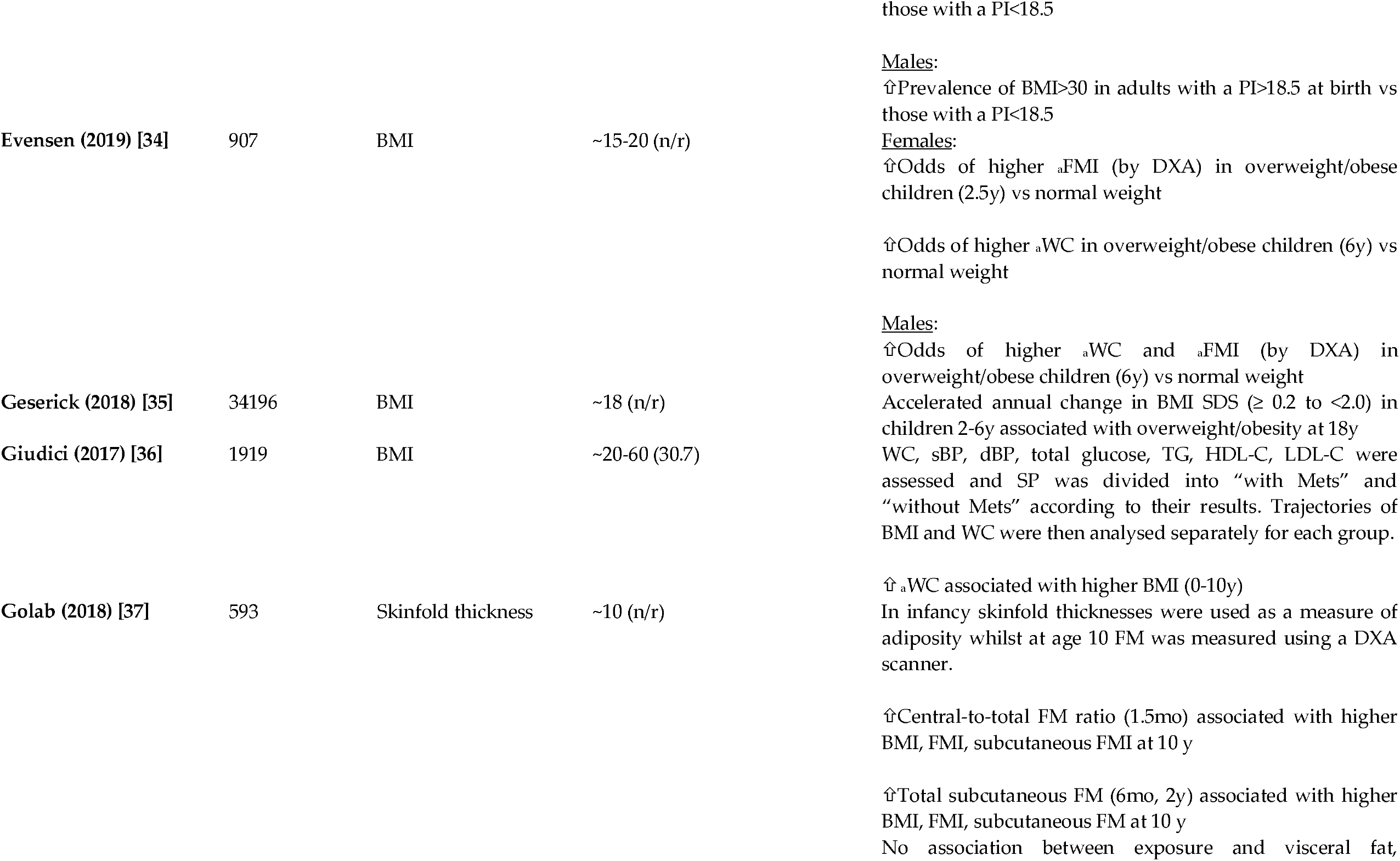
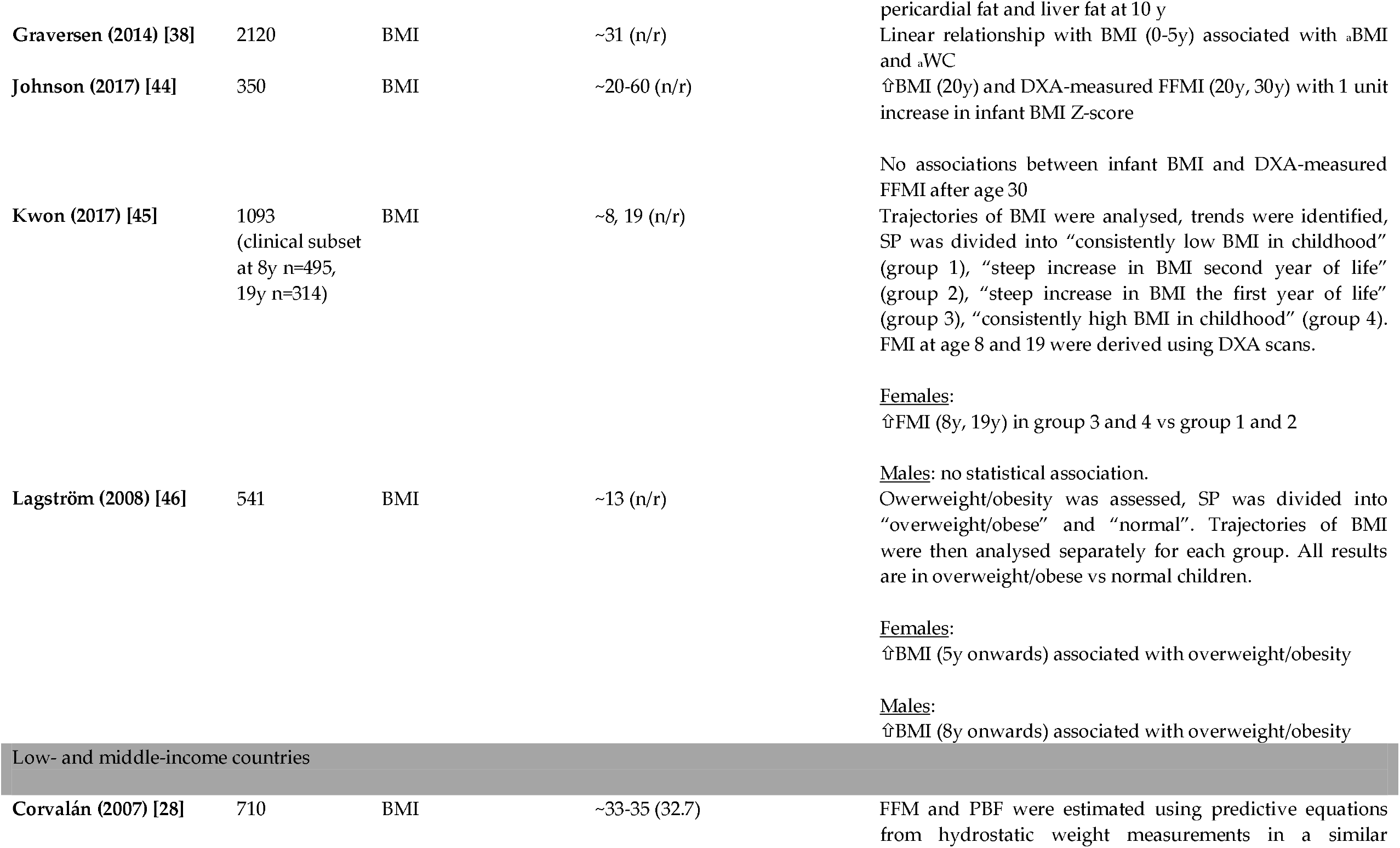
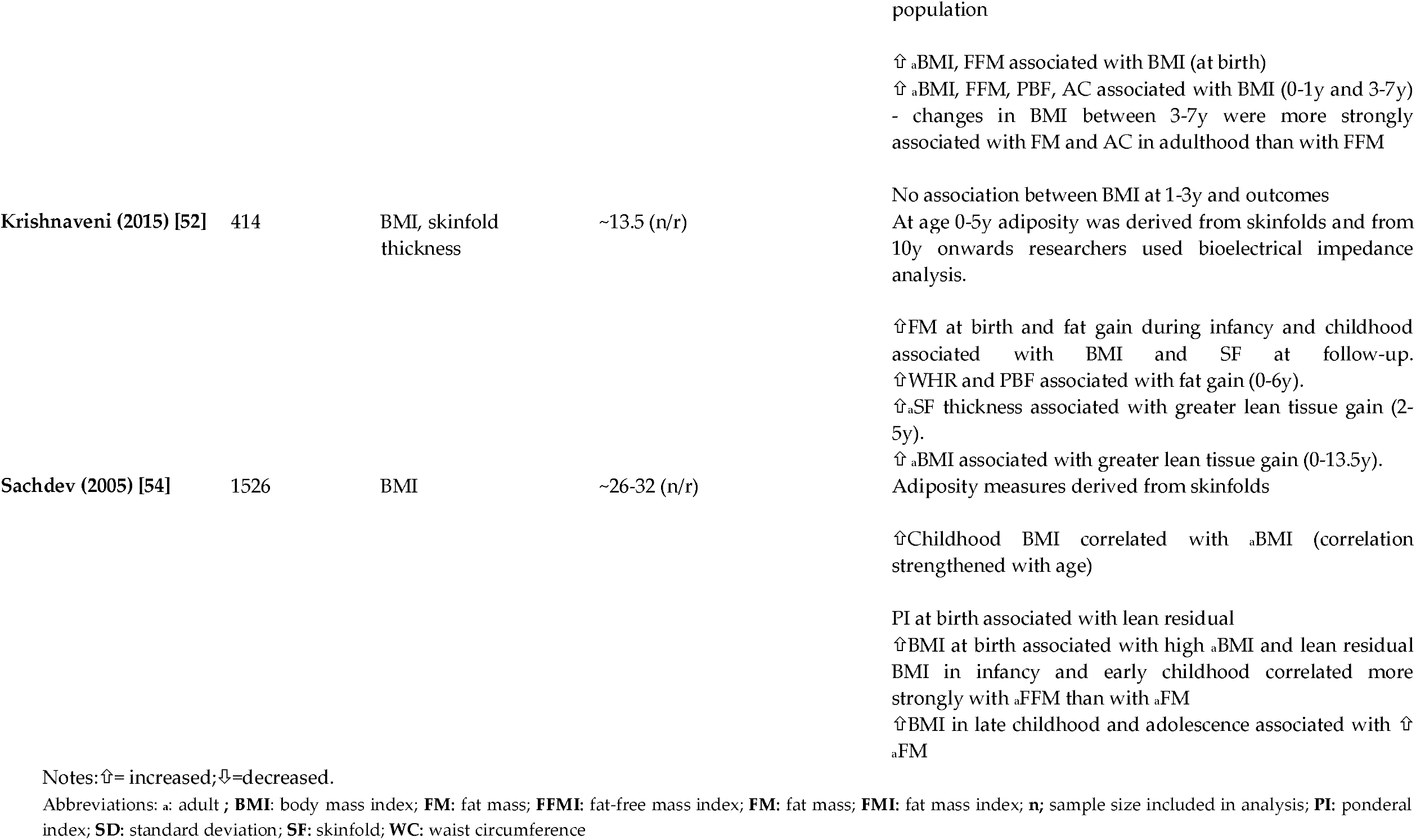
Summary of studies reporting on obesity-related outcomes.

#### 3.3.1 Cardiovascular outcomes

##### High-income countries

A study looking at cardiovascular disease (CVD) risk found that females who had increased BMI and skinfold (SF) thicknesses from ages 1-5y had increased risk of CVD (BMI: p<0.001; SF thicknesses: p<0.05). High-risk males had increased BMI at 3y (p<0.001) and increased SF thicknesses from 3-5y (p<0.001) [42]. Another study reporting on CVD risk did not find any association before 5y, but reported that increased BMI in later childhood was associated with increased CVD risk [40].

Four studies reported on blood pressure (BP). One found that low BMI at birth was associated with increased BP. The researchers also found that there was no association between BMI below 7y and later BP, however, those subjects with the highest BP had a low birthweight and were overweight or obese at age 2 and time of exposure [41]. Another reported that a 1 unit increase in BMI SDS from birth to 7y was associated with elevated blood pressure. Changes in systolic blood pressure (sBP) were greater in females than in males (1.4 mmHg vs 0.7mmHGg), but in contrast diastolic blood pressure (dBP) was greater in males than in females (1.0mmHg vs 0.5mmHg) [39]. Another study measuring BP reported that children who became obese early in life and who had a delayed overweight (overweight at 10 and 18y) had higher BP at follow-up than those with a healthy weight in childhood (all p<0.001) [49]. A Finnish study found that sBP fell both with increasing birth weight and increasing BMI at 2y. The researchers also reported on the prevalence of coronary events and found that adults who experienced coronary events were smaller than average at birth and had a BMI below average at age 2. After age 2 and 4 (for boys and girls, respectively), their BMI increased progressively. The authors concluded that “The risk of coronary events was more strongly related to the tempo of childhood gain in BMI than to the BMI attained at any particular age” [30].

Results from a British cohort examined carotid intima-media thickness (cIMT) and found that boys with a BMI in the upper quartiles had increased odds of high cIMT with 1 unit increase in z-score BMI at 4y (OR1.26;p=0.03) vs boys with a BMI in the three lower quartiles. They found no such association in women (p>0.05) [56].

Another British cohort reported on lipid levels at age 53 and found that a 1 SD increase in BMI at ages 2 and 4 was associated with lower levels of total cholesterol (TC) (p=0.007 and p=0.003 respectively) and an increase in BMI from 7-15y was associated with lower levels of high-density lipoprotein cholesterol (HDL-C) with the association being stronger and greater in women. The researchers adjusted for current body size [48].

##### Low- and middle-income countries

A study from India found that fat gain measured by skinfolds from 5y onwards was associated with elevated sBP in adulthood [52].

A study from Melanesia reporting on TC found that BMI z residuals from 0-5y in males were associated with increased TC. In females, there was an positive association between skinfold (SF) residuals from 0-5y and CVD, and BMI z residuals from 6-11y and CVD [55].

#### 3.3.2 Glucose metabolism outcomes

##### High-income countries

Two studies reported on the risk of type 2 diabetes (T2D). One from Finland found that a gain of 1kg/m2 in subjects who had the minimum BMI from 3-11y had an increased risk of T2D vs those who gained less than 1kg/m2 (OR1.87;p=0.04) [47]. Another Finnish study reported that children who had a BMI above the study population median at 11y had a decreased risk of T2D with each unit increase in BMI z-score from 0-7 y. After 7y the same group had an increased risk of T2D with each unit increase in BMI z-score. Association was greater and stronger in females (females: OR1.35;p=0.004, males: OR1.23;p=0.01). Researchers found that within the same group, a low BMI at birth and at 2y and a high BMI at 11y were associated with T2D. The group with a BMI below the study population median at 11y had a decreased risk of T2D with each unit increase in BMI z-score from 0-11y. In this group a low BMI at birth was associated with T2D [33].

A different Finnish study assessed fasting plasma glucose and insulin resistance and found that low birth weight, low BMI at 2y and an increase in SD scores for BMI from 2-11y were associated with raised fasting plasma glucose and insulin resistance [30].

A study reporting on hyperglycaemia in Chile found that subjects with hyperglycaemia typically had an increased BMI from approximately 2y onwards [31].

##### Low- and middle-income countries

A study from India looking at impaired glucose tolerance/diabetes mellitus (IGT/DM) found that a PI at birth and BMI in childhood close to a z-score of 0 were protective against IGT/DM (OR0.80;p=0.04 and OR0.77;p<0.001, respectively). Greater changes in BMI z-score from birth to adulthood were associated with increased odds of IGT/DM that grew stronger by age (all p<0.001, see Appendix B for all ORs). Researchers concluded that those with IGT/DM in adulthood were typically LBW-infants and that IGT/DM was associated with low BMI in childhood, followed by an accelerated BMI gain between birth, infancy, childhood or adolescence and adulthood [53].

Another study from India reporting on DM found that there were increased odds of diabetes per SD score BMI increase from 2-11y (OR1.25;p=0.01) and that subjects with diabetes had more rapid weight/BMI gain throughout infancy, childhood and adolescence as well as a lower BMI in infancy [51]. A different study, using the study population from the same birth cohort, found a similar association: increased odds of IGT/DM with a low BMI at 1y and with a 1SD BMI z-score increase from 2-12y, which attenuated after adjustment for current body size (OR1.36;p<0.001 and OR1.26;p=0.004, respectively). Researchers also found an association with low PI at birth and increased plasma glucose and insulin concentrations and insulin resistance at follow-up, and noted that subjects who developed DM/IGT typically had a lower PI and BMI up to the age of 2 [50]. A study reporting on insulin concentrations did not find an association with BMI and skinfolds in the first 5y post-natal but did associate fat gain measured by skinfolds from 5y onwards with increased fasting insulin concentrations and insulin resistance [52].

One study reporting on total glucose did not find any association with BMI and skinfolds in early life but did find that BMI residuals in females aged 12-19y were associated with increased total glucose. In males, the researchers reported that SF residuals at ages 6-11y and 12-19y were associated with increased levels of total glucose [55].

#### 3.3.3 Metabolic syndrome outcomes

##### High-income countries

Four studies reported on metabolic syndrome (MetS). One study found no association with MetS and body size at birth and 2 years. However, changes in BMI in infancy were predictive, with a 1 SD z-score increase in BMI from 0-2y in males was associated with decreased odds of MetS in adulthood (OR0.53;0.33-0.87). Though similar was observed in women, those changes were not statistically significant. Researchers adjusted for current body size and did not report unadjusted results [27].

Another study reported that subjects with a BMI in the ≥50-<75 and ≥95 percentile had increased risk of MetS vs those in the ≥5-<50 percentile (RR1.9 vs RR1.6). Subjects with a BMI above the ≥95 percentile at 4y and 5y had a slightly greater risk of MetS vs those in the ≥5-<50 percentile (RR2.5 vs RR2.4) [38]. Similarly, another study from France found that subjects with MetS had an increased BMI at 4-6y and 7-10y (p=0.01 and p<0.001, respectively) [36].

A Chilean study reported that subjects who had MetS, had higher and faster growth in BMI from ages 6mo-23y [31].

##### Low- and middle-income countries

Fall et al. found that the odds of MetS increased per SD score BMI change from 2-11y (OR1.48; p<0.001) and that subjects with MetS had a more rapid weight/BMI gain throughout infancy, childhood and adolescence [51].

#### 3.3.4 Obesity-related outcomes

##### High-income countries

11 studies reported on obesity-related outcomes. A study reporting on BMI did not find any association before the age of 5. BMI from 5y onwards was associated with overweight/obesity in females whilst this association in males was seen from age 8y onwards [46].

Angbratt et al. found a small correlation with PI at birth, 1.5y, and BMI at 2.5y (r<0.5) and overweight/obesity at follow-up whilst BMI at age 5, 7 and 10y was strongly correlated with BMI at follow-up (r>0.5) [29]. Another study found that a linear relationship with BMI at ages 0-5y was associated with higher BMI and waist circumference (WC) at follow-up [38].

One study reported that an increased BMI from 0-10y was associated with elevated WC at follow-up (p<0.001) [36]. Similarly, a study reported that obese subjects at follow-up had a high BMI between 0-12y (all p<0.001). The researchers also found that females and males with a PI<18.5 at birth had increased odds of becoming obese adults vs those with a PI>18.5 (OR3 and OR4, respectively) [32]. Similarly, another study found that subjects with abdominal obesity had an increased BMI gain from infancy to follow-up versus those without abdominal obesity[31].

A study reporting on accelerated annual change in BMI SDS found that an annual change of ≥ 0.2 to <2.0 BMI SDS in children 2-6y increased their risk of overweight/obesity later in life vs children with a stable BMI between age 2-6y (RR1.43; CI1.35-1.49) [35].

One study reported on infant BMI only and found that a 1 unit increase in infant BMI z-score was associated with high BMI at 20y (β=0.70; CI0.31-1.09; p<0.001) and high DXA-measured fat-free mass index (FFMI) at 20y (β=0.75; CI0.37-1.12;p<0.001) and 30y (β=0.34; CI0.12-0.56). They found no association between infant BMI and body composition after age 30 [44].

Golab et al. reported on different adiposity measures using skinfolds and found that an increased central-to-total fat mass (FM) ratio at 1.5mo and increased total subcutaneous FM at 6 months and 2 y was associated with higher BMI and fat mass index (FMI) at follow-up [37]. Similarly, another study reported that females who had a steep increase in BMI the first year of life had higher DXA-measured FMI at follow-up. There was no association in males [45].

The Norwegian study showed that females who were overweight/obese at 2.5y had increased odds of a higher FMI measured by DXA at follow-up vs those with a normal weight at 2.5y (OR:2.00; p<0.05), however, the association was stronger with overweight/obesity at 6y and increased WC at follow-up vs normal weight at 6y (OR:4.79; p<0.001). In males there was no association between overweight/obesity at age 2.5 and obesity-related outcomes at follow up; however overweight/obesity at 6y was associated with increased odds of increased WC (OR:5.56;p<0.001) and DXA-measured FMI (OR:4.14; p<0.001) at follow-up [34].

##### Low- and middle-income countries

A Guatemalan study found an association between BMI at birth and BMI (β=0.33;p<0.05) and fat-free mass (FFM) (β=0.49;p<0.01) at follow-up. FFM was estimated using predictive equations from hydrostatic weight measurements in a similar population. Results also showed that BMI at 0-1y and 3-7y was associated with BMI, FFM, percentage body fat (PBF) and abdominal circumference (AC), and that changes in BMI from 3-7y were most strongly associated with adult FM and AC (see Appendix B for details). There was no association between BMI at ages 1-3y and measured outcomes at follow-up [28].

A similar study in India found a correlation between BMI in childhood and BMI at follow-up; correlation strengthened with age (6mo: r=0.19, 2y: r=0.24, 5y: r= 0.32, 14y: r=0.65). The study reported that PI at birth was associated with FFM in adulthood and that BMI in infancy and early childhood correlated more strongly with adult FFM whilst increased BMI in late childhood and adolescence was associated with adult FM. FFM and FM were derived from skinfolds [54].

Another Indian study reported an association between fat gain from 0-6y and waist-hip ratio, BMI and PBF at follow-up. In addition, the results showed that greater lean tissue gain from 2-5y and 0-13.5y was associated with SF thickness and BMI at follow-up, respectively. FM, FFM and PBF were derived from skinfolds up to age 5 whereafter researchers used bioelectrical impedance analysis [52].

## 4. Discussion

### 4.1 Summary

The major finding from our review is that evidence on childhood body composition and later life NCD is severely limited. Though four studies assessed skinfold thickness in childhood, we did not find any using the more direct and technically superior methods such as isotope dilution, plethysmography or X-ray. We did however find numerous studies using BMI (and a smaller number using Ponderal Index) - but it is important to note that these are only proxy measures of body composition. Even with BMI as the childhood exposure variable, associations with later NCD are difficult to interpret due to marked inter-study heterogeneity, especially in terms of NCD measure and age at follow-up. Varied approaches to analysing, reporting and presenting data add to the challenge of interpreting what is. Most studies showed that childhood BMI is associated with later-life cardiometabolic NCD risk and that changes in BMI rather than absolute BMI, appears to be important. Some studies also showed sex-specific differences. Most studies were unadjusted for current body size and thus the independent effect of childhood BMI is open to question. Because most studies were from high-income settings, wider generalizability to populations in low- and middle-income settings is unknown.

### 4.2 Interpretation of findings

Most of our interpretable data uses BMI as the childhood exposure variable. BMI is widely used to categorize nutritional status because it is simple and can be compared with reference standards [57]. Many people and even many non-specialist scientists/clinicians also view it as “an indicator of body fatness”[58] – hence why it was so common in our search results. It is however just an indicator of excess or inadequate weight, not excess or inadequate fat mass (FM) and it cannot differentiate between FM and fat-free mass (FFM). The perception about “fatness” arises because of an association between BMI and FM in adulthood especially, but in children, this is influenced by variety of other factors such as age, sex, pubertal status, and ethnicity. In relation to using BMI as an indicator of bodyfat in early life, another big limitation arises: specifically, that low BMI at birth and during infancy can act as a proxy for low FFM [8], and hence as a marker of poor capacity for metabolic homeostasis [59]. This is highly problematic as this implies that greater relative weight indexes different things at different time points. Although several studies in this review have shown that infant weight gain is protective of NCDs in later life and that both low BMI at birth and in infancy *and* high childhood BMI is associated with an increased risk of NCDs, the lack of information about the relationship between BMI and body composition makes it difficult to interpret the data and establish clear associations. Infancy is a particularly challenging period to investigate, as low BMI may indicate low FFM, whereas rapid BMI increases over time may indicate fat deposition through catch-up growth [59].

In the studies that did differentiate between FM and FFM, low BMI in infancy and high BMI in childhood both predicted later NCD risk. This pattern links with the ‘capacity-load model’ which hypothesizes that increased size at birth indicates a greater metabolic (homoeostatic) capacity, although those born in the highest weight-categories may deviate from this pattern since a higher proportion of their weight is likely due to adipose tissue, which imposes a metabolic load [18,60].

Several studies have associated birth weight and BMI in infancy with adult FFM, whilst BMI in later childhood was associated with both FM and FFM [61–63]. This is consistent with studies included in this review. Four studies found that early BMI was associated with adult FFM whilst BMI in later childhood was associated with FM and FFM [28,34,64,65]. These were also the only studies distinguishing between FM and FFM while the rest of the studies reported on BMI as a whole. As BMI does not distinguish between FM and FFM, associations between early BMI and overweight/obesity are likely to be confounded by the gain in FFM, thus are a threat to the validity of its use. This might also explain, why some studies did find an association between BMI and NCDs, while others did not.

Other inconsistencies of our results among studies reporting on the same outcome measure can be explained by the follow-up time of the respective studies. In our review, years of follow-up varied widely from 10-70y. In studies with relatively short follow-up time that show no association between exposure and outcome, it is likely that some subjects will go on to develop a NCD with time as most NCDs do not develop until later in life [66–68]. T2D, for instance, is most commonly seen in people over the age of 45 [68], and only 1/3 studies in current review with T2D as an outcome, had a study population above age 45 [33].

Lack of adjusting for current body size also impacts interpretation of our findings. In 1999, Lucas et al. criticized researchers’ lack of understanding and communication of the statistical implications of this. Over 20 years later, our review suggests that the problem remains. Adjustment for current size is important because it implies that change in size as well as initial size can contribute to an association [69]. Previous reviews found that studies which had adjusted for current body size experienced a partial attenuation in effect size [70] and that some associations completely disappeared after adjustment for current body size [71]. These discoveries show that studies that fail to undertake these adjustments may be confounded by adult body size, and therefore the observed associations might in fact reflect the tracking of childhood BMI across the lifespan instead of an actual association [72].

In our review, 11 of 29 studies adjusted for current body size [27,30,56,40,41,44,48–51,53] and like previously reported, some associations attenuated or became statistically insignificant after adjustments or even reversed. However, while the researchers did comment on the effect adjustment for body size had on the results, most of the studies did not report both adjusted vs unadjusted results in respect to adjustment for current body size, making readers unable to analyse and interpret raw data to draw their own conclusions.

### 4.3 Research in context

Similar to our review, Park et al. found an association between childhood overweight (2-12y), unadjusted for adult body size, and CVD outcomes in adulthood. They were unable to conclude that childhood overweight is an independent risk factor of adult CVD as the few studies that did report adjusted results, were inconclusive. Furthermore, studies were mainly from high-income settings and thus the generalizability is limited [24]. In contrast, a review from Owen et al. in 2009 concluded, that BMI gain from age 2-6 had a weak inverse association (RR0.94, 95%CI:0.82-1.07) with CHD risk [73]; however statistical findings are weak with crossing CIs. It is also important to notice that this conclusion was based on only 3 estimates and that the researchers for this review did not exclude cohorts with high-risk subjects (e.g. LBW babies). Owen et al. also reported that the inverse association between childhood BMI and CHD risk became weakly positive after age 7y and grew stronger with age [73]. The inverse association is consistent with some studies included in the current review that found an association between low BMI in infancy and NCD risk factors in adulthood. The evidence supports the capacity-load model hypothesis [74], where LBW means lower capacity, but an excessively high birthweight, likewise, also means lower capacity in terms of ability of metabolically coping with NCDs. Results from a number of studies in the current review suggest the same trend but whether this is due to the uncontrolled adjustments for current body size remains unanswered.

A review by Simmonds et al. based on HIC studies, reported that BMI has poor sensitivity in identifying healthy-weight children, who later would become obese adults. However, BMI was found to be a reasonable accurate measure of obesity and thus can identify obese children who most likely will become obese adults. The researchers also reported that obese children had more than five times the risk of becoming obese adults than non-obese children (RR5.21; 95%CI4.50-6.02) [21]. These findings are consistent with studies we found, which suggest a pattern whereby increased BMI at different ages throughout childhood is associated with NCD/obesity-related outcomes in adult life. Simmonds et al. also found that BMI was a poor predictor of obesity-related diseases, as only 40% of adult diabetes and 20% of CHD would occur in overweight/obese children [21]. This further underlines the importance of using better body composition measurements in future studies, to examine the effect of childhood fat mass and fat-free mass on adult NCDs.

These three reviews also experienced challenges with the diversity in reporting, which for Simmonds et al. meant that a number of assumptions were made to conduct the meta-analysis and thus the reliability of the pooled estimate may be limited [21]. Due to the limitations of these review, results should be interpreted as a general trend rather than a precise estimate of an association or predictive accuracy.

Finally, a 2021 review focused on NCD risk in survivors of childhood *under*nutrition/famine [15]. Though the exposure was to undernutrition (as assessed by standard anthropometric measures) and thus the opposite type of malnutrition to most studies in this review, authors also found an association with numerous NCD-related outcomes. Strength and consistency of association also varied according to outcome. Interpreting the reviews together, it seems that extremes at both ends of the malnutrition spectrum risk long term adverse outcomes. Our observation that rate of weight change can mediate risk might offer insights into mechanism spanning the two types of malnutrition. As that review highlights in the conclusions, this work on mechanisms is urgently needed.

### 4.4 Limitations

All included studies controlled for some known confounders. However, all studies were also of observational design, and there is therefore an inherent risk of residual confounding affecting the results. Whilst it is impossible to control for all confounders, the most evident and important confounders should be taken into consideration. For example, Bhargava et al. did not adjust for socio-economic status (SES) [50]. SES is a well-known confounder and lack of controlling thereof may lead to significantly affected and incorrect effect size [75]. Though not simple, it might also have been possible to control for different times of follow-up e.g. using age-standardized reporting of NCD-related outcome measures.

As mentioned, very few studies reported on actual body composition in relation to NCDs which consequently highly limits our understanding of how FM and FFM in early childhood relates to later NCD risk. Alongside the problem of using BMI rather than other true measures of body composition, adjustment for current body size was a major limitation in this review. Since under half of the studies included in the narrative synthesis adjusted for current size, we are unable to confirm the independent effect of early childhood body size on long-term cardiometabolic health, and thus there is a possibility that the associations seen in studies that failed to undertake these adjustments, is mediated through adult body size.

Several studies were greatly affected by loss to follow-up and only four had an attrition rate below 20% [28,34,49,52]. Seven studies reported a loss to follow-up above 60% of the original cohort [40–42,48,53,55,65], while eight did not address attrition rate at all nor did they report it [29,30,32,33,45–47,56].

None of the studies presented power calculations for their sample size, and only four studies [31,41,47,53] identified reduced power as a limitation of their study and possible explanation for the lack of weak association/difference in groups.

## 5. Research recommendations

This review has highlighted several areas needing urgent research attention.

Heterogeneity among future studies might be reduced by researchers reading our review when planning their own work and choosing outcome variables / measurement timings which can then be more directly compared with this past work. Checklists of key items to report in such nutrition/NCD follow-up studies might also help, forming the basis for a STROBE checklist extension [76].

Based on the risk of bias assessment, it is recommended that future longitudinal studies improve their reporting on several potential sources of bias and include a flow diagram to demonstrate their participation and response rates. In particular, follow-up rates should be reported as well as implications clearly discussed.

BMI is a poor measure of adiposity as it does not distinguish between FM and FFM. Future work should use additional, more direct measures of adiposity, e.g. peapod, isotope dilution, DXA. These studies urgently needed and could offer valuable insights into mechanisms linking early life malnutrition (both undernutrition and overweight/obesity) with later life NCD risk. Studies are also needed to explore the relative utility of different methods: e.g. which field-appropriate measures (such as BIA) are most closely associated with the more complex, costly but arguably more ‘gold standards’ measures such as DEXA scans.

Finally, less than half of studies in this review adjusted for current body size. Future studies should present both crude and adjusted associations.

## 6. Conclusion

Our review found that early life nutritional status, mostly as assessed by low BMI in infancy and increased BMI in later childhood, was often associated with increased risk of cardiometabolic diseases and risk factors in adult life. Although exact patterns of association varied in different studies and settings (i.e. whether absolute BMI or BMI change in childhood matter most), some evidence in our review suggest a pattern where low BMI at birth and infancy followed by a rapid weight gain in childhood exceeding recommended levels increases the risk of NCDs. Whether different patterns of body composition mediate or explain some of these variations is not possible to say. Neither is it known whether childhood BMI is an independent risk factor for NCDs in adulthood, or whether the association simply is mediated through adult overweight/obesity. Due to the limited evidence on nutritional status measures other than BMI, it is not possible to identify which measure of body composition best predicts NCDs in adulthood. We highlight several gaps in literature: high quality evidence on this topic - in particular evidence from low- and middle-income countries and; the use of more direct measures of body composition to better describe nutritional status. As technology is rapidly improving, better equipment/solutions are more accessible and can provide research with adequate measures of body composition. Findings from our review underline the necessity to improve and continue the tracking of body composition from birth to adulthood to help understand relevant mechanisms linking child nutrition to adult health/NCD. This has a key role to play in preventing the increasing rates of overweight/obesity among children and adults and ultimately prevent the rising prevalence of NCDs.

## Data Availability

All data referred to in the manuscript are available on the electronic databases that were searched in this review.

## Supplementary Materials

The following are available online at www.mdpi.com/xxx/s1: None.

## Author Contributions

Conceptualization, M.K. and A.B.; methodology, M.K. and A.B.; formal analysis, A.B.; investigation, A.B. and I.G.; resources, writing—original draft preparation, A.B.; writing—review and editing, A.B., M.K., J.C.K.W., A.J.M., V.O.O., C.U.L. visualization, A.B.; supervision, M.K.; project administration, A.B.; funding acquisition, M.K. All authors have read and agreed to the published version of the manuscript. Please turn to the <u>CRediT taxonomy</u> for the term explanation. Authorship must be limited to those who have contributed substantially to the work reported.

## Acknowledgments

In this section you can acknowledge any support given which is not covered by the author contribution or funding sections. This may include administrative and technical support, or donations in kind (e.g., materials used for experiments).

## Appendix A

### Full search strategy

Database: Ovid MEDLINE(R) and In-Process & Other Non-Indexed Citations and Daily <1946 to July 27, 2020> Search Strategy:

--------------------------------------------------------------------------------

1. (diabetes type 2 or type 2 diabetes or diabetes mellitus or insulin resistance syndrome* or insulin resistan* or hyperglycaemia or hypertension or arteriosclerosis or cardiovascular disease* or cardio vascular disease* or blood pressure or coronary heart disease* or metabolic syndrome* or dysmetabolic syndrome* or cardio metabolic disorder* or cardiometabolic disorder* or lipid profile or lipid metabolism or lipid* profile or glucose metaboli* disorder* or glucose intoleran* or glucose toleran* or obes* or artheroscleros*).mp. (1781365)
2. ((infan* or baby or babies or child* preschool or preschool child or early childhood or young children or kindergarten* or children under 5 or children under five or under 5’s) adj10 (body composition or body fat or fat percentage or fat % or lean mass or fat mass of fat free mass or muscle mass or grip strength or hand strength or antropometr* or skinfold or skinfold thickness or body mass index or BMI or percent fat or fat percent or densitometry)).mp. (5242)
3. 1 and 2 (2221)
4. ((infan* or baby or babies or child* preschool or preschool child or early childhood or young children or kindergarten* or children under 5 or children under five or under 5’s) adj10 (body composition or body fat or fat percentage or fat % or lean mass or fat mass of fat free mass or muscle mass or grip strength or hand strength or antropometr* or skinfold or skinfold thickness or body mass index or BMI)).mp. [mp=title, abstract, original title, name of substance word, subject heading word, floating sub-heading word, keyword heading word, organism supplementary concept word, protocol supplementary concept word, rare disease supplementary concept word, unique identifier, synonyms] (5194)
5. 1 and 4 (2219)
6. body <u>composition.mp</u>. [mp=title, abstract, original title, name of substance word, subject heading word, floating sub-heading word, keyword heading word, organism supplementary concept word, protocol supplementary concept word, rare disease supplementary concept word, unique identifier, synonyms] (57816)
7. body <u>fat.mp</u>. [mp=title, abstract, original title, name of substance word, subject heading word, floating sub-heading word, keyword heading word, organism supplementary concept word, protocol supplementary concept word, rare disease supplementary concept word, unique identifier, synonyms] (33004)
8. (fat percentage or fat %).mp. [mp=title, abstract, original title, name of substance word, subject heading word, floating sub-heading word, keyword heading word, organism supplementary concept word, protocol supplementary concept word, rare disease supplementary concept word, unique identifier, synonyms] (271026)
9. fat <u>mass.mp</u>. [mp=title, abstract, original title, name of substance word, subject heading word, floating sub-heading word, keyword heading word, organism supplementary concept word, protocol supplementary concept word, rare disease supplementary concept word, unique identifier, synonyms] (20698)
10. lean <u>mass.mp</u>. [mp=title, abstract, original title, name of substance word, subject heading word, floating sub-heading word, keyword heading word, organism supplementary concept word, protocol supplementary concept word, rare disease supplementary concept word, unique identifier, synonyms] (5400)
11. fat free <u>mass.mp</u>. [mp=title, abstract, original title, name of substance word, subject heading word, floating sub-heading word, keyword heading word, organism supplementary concept word, protocol supplementary concept word, rare disease supplementary concept word, unique identifier, synonyms] (7537)
12. muscle <u>mass.mp</u>. [mp=title, abstract, original title, name of substance word, subject heading word, floating sub-heading word, keyword heading word, organism supplementary concept word, protocol supplementary concept word, rare disease supplementary concept word, unique identifier, synonyms] (17698)
13. grip <u>strength.mp</u>. [mp=title, abstract, original title, name of substance word, subject heading word, floating sub-heading word, keyword heading word, organism supplementary concept word, protocol supplementary concept word, rare disease supplementary concept word, unique identifier, synonyms] (11357)
14. hand <u>strength.mp</u>. [mp=title, abstract, original title, name of substance word, subject heading word, floating sub-heading word, keyword heading word, organism supplementary concept word, protocol supplementary concept word, rare disease supplementary concept word, unique identifier, synonyms] (15132)
15. anthropometr*.mp. [mp=title, abstract, original title, name of substance word, subject heading word, floating sub-heading word, keyword heading word, organism supplementary concept word, protocol supplementary concept word, rare disease supplementary concept word, unique identifier, synonyms] (78466)
16. (skinfold or skin fold thickness).mp. [mp=title, abstract, original title, name of substance word, subject heading word, floating sub-heading word, keyword heading word, organism supplementary concept word, protocol supplementary concept word, rare disease supplementary concept word, unique identifier, synonyms] (10959)
17. (body mass index or BMI).mp. [mp=title, abstract, original title, name of substance word, subject heading word, floating sub-heading word, keyword heading word, organism supplementary concept word, protocol supplementary concept word, rare disease supplementary concept word, unique identifier, synonyms] (279689)
18. 6 or 7 or 8 or 9 or 10 or 11 or 12 or 13 or 14 or 15 or 16 or 17 (608903)
19. (infan* adj10 #13).mp. [mp=title, abstract, original title, name of substance word, subject heading word, floating sub-heading word, keyword heading word, organism supplementary concept word, protocol supplementary concept word, rare disease supplementary concept word, unique identifier, synonyms] (4182)
20. ((baby or babies) adj10 #13).mp. [mp=title, abstract, original title, name of substance word, subject heading word, floating sub-heading word, keyword heading word, organism supplementary concept word, protocol supplementary concept word, rare disease supplementary concept word, unique identifier, synonyms] (551)
21. ((child* preschool or preschool child) adj10 #13).mp. [mp=title, abstract, original title, name of substance word, subject heading word, floating sub-heading word, keyword heading word, organism supplementary concept word, protocol supplementary concept word, rare disease supplementary concept word, unique identifier, synonyms] (2)
22. (early childhood adj10 #13).mp. [mp=title, abstract, original title, name of substance word, subject heading word, floating sub-heading word, keyword heading word, organism supplementary concept word, protocol supplementary concept word, rare disease supplementary concept word, unique identifier, synonyms] (60)
23. (young children adj10 #13).mp. [mp=title, abstract, original title, name of substance word, subject heading word, floating sub-heading word, keyword heading word, organism supplementary concept word, protocol supplementary concept word, rare disease supplementary concept word, unique identifier, synonyms] (91)
24. (kindergarten adj10 #13).mp. [mp=title, abstract, original title, name of substance word, subject heading word, floating sub-heading word, keyword heading word, organism supplementary concept word, protocol supplementary concept word, rare disease supplementary concept word, unique identifier, synonyms] (50)
25. ((children under 5 or children under five or under 5’s) adj10 #13).mp. [mp=title, abstract, original title, name of substance word, subject heading word, floating sub-heading word, keyword heading word, organism supplementary concept word, protocol supplementary concept word, rare disease supplementary concept word, unique identifier, synonyms] (66)
26. 19 or 20 or 21 or 22 or 23 or 24 or 25 (4931)
27. (diabetes type 2 or type 2 diabetes or diabetes mellitus or insulin resistance syndrome* or insulin resistan* or hyperglycaemia or hypertension or arteriosclerosis or cardiovascular disease* or cardio vascular disease* or blood pressure or coronary heart disease* or metabolic syndrome* or dysmetabolic syndrome* or cardio metabolic disorder* or cardiometabolic disorder* or lipid profile or lipid metabolism or lipid* profile or glucose metaboli* disorder* or glucose intoleran* or glucose toleran* or obes* or hypertension or artheroscleros*).mp. (1781365)
28. (diabetes type 2 or type 2 diabetes).mp. [mp=title, abstract, original title, name of substance word, subject heading word, floating sub-heading word, keyword heading word, organism supplementary concept word, protocol supplementary concept word, rare disease supplementary concept word, unique identifier, synonyms] (126496)
29. diabetes <u>mellitus.mp</u>. [mp=title, abstract, original title, name of substance word, subject heading word, floating sub-heading word, keyword heading word, organism supplementary concept word, protocol supplementary concept word, rare disease supplementary concept word, unique identifier, synonyms] (433938)
30. insulin resistance syndrome*.mp. [mp=title, abstract, original title, name of substance word, subject heading word, floating sub-heading word, keyword heading word, organism supplementary concept word, protocol supplementary concept word, rare disease supplementary concept word, unique identifier, synonyms] (1746)
31. insulin resistan*.mp. [mp=title, abstract, original title, name of substance word, subject heading word, floating sub-heading word, keyword heading word, organism supplementary concept word, protocol supplementary concept word, rare disease supplementary concept word, unique identifier, synonyms] (98055)
32. <u>hyperglycaemia.mp</u>. [mp=title, abstract, original title, name of substance word, subject heading word, floating sub-heading word, keyword heading word, organism supplementary concept word, protocol supplementary concept word, rare disease supplementary concept word, unique identifier, synonyms] (10223)
33. <u>.hypertension.mp</u>. [mp=title, abstract, original title, name of substance word, subject heading word, floating sub-heading word, keyword heading word, organism supplementary concept word, protocol supplementary concept word, rare disease supplementary concept word, unique identifier, synonyms] (485390) 34.
34. <u>arteriosclerosis.mp</u>. [mp=title, abstract, original title, name of substance word, subject heading word, floating sub-heading word, keyword heading word, organism supplementary concept word, protocol supplementary concept word, rare disease supplementary concept word, unique identifier, synonyms] (70980)
35. cardiovascular disease*.mp. [mp=title, abstract, original title, name of substance word, subject heading word, floating sub-heading word, keyword heading word, organism supplementary concept word, protocol supplementary concept word, rare disease supplementary concept word, unique identifier, synonyms] (260492)
36. cardio vascular disease*.mp. [mp=title, abstract, original title, name of substance word, subject heading word, floating sub-heading word, keyword heading word, organism supplementary concept word, protocol supplementary concept word, rare disease supplementary concept word, unique identifier, synonyms] (705)
37. blood <u>pressure.mp</u>. [mp=title, abstract, original title, name of substance word, subject heading word, floating sub-heading word, keyword heading word, organism supplementary concept word, protocol supplementary concept word, rare disease supplementary concept word, unique identifier, synonyms] (442915)
38. coronary heart disease*.mp. [mp=title, abstract, original title, name of substance word, subject heading word, floating sub-heading word, keyword heading word, organism supplementary concept word, protocol supplementary concept word, rare disease supplementary concept word, unique identifier, synonyms] (50273)
39. metabolic syndrome*.mp. [mp=title, abstract, original title, name of substance word, subject heading word, floating sub-heading word, keyword heading word, organism supplementary concept word, protocol supplementary concept word, rare disease supplementary concept word, unique identifier, synonyms] (57036)
40. dysmetabolic syndrome*.mp. [mp=title, abstract, original title, name of substance word, subject heading word, floating sub-heading word, keyword heading word, organism supplementary concept word, protocol supplementary concept word, rare disease supplementary concept word, unique identifier, synonyms] (111)
41. cardio metabolic disorder*.mp. [mp=title, abstract, original title, name of substance word, subject heading word, floating sub-heading word, keyword heading word, organism supplementary concept word, protocol supplementary concept word, rare disease supplementary concept word, unique identifier, synonyms] (87)
42. cardiometabolic disorder*.mp. [mp=title, abstract, original title, name of substance word, subject heading word, floating sub-heading word, keyword heading word, organism supplementary concept word, protocol supplementary concept word, rare disease supplementary concept word, unique identifier, synonyms] (562)
43. lipid* <u>profile.mp</u>. [mp=title, abstract, original title, name of substance word, subject heading word, floating sub-heading word, keyword heading word, organism supplementary concept word, protocol supplementary concept word, rare disease supplementary concept word, unique identifier, synonyms] (22957)
44. lipid <u>metabolism.mp</u>. [mp=title, abstract, original title, name of substance word, subject heading word, floating sub-heading word, keyword heading word, organism supplementary concept word, protocol supplementary concept word, rare disease supplementary concept word, unique identifier, synonyms] (99667)
45. glucose metaboli* disorder*.mp. [mp=title, abstract, original title, name of substance word, subject heading word, floating sub-heading word, keyword heading word, organism supplementary concept word, protocol supplementary concept word, rare disease supplementary concept word, unique identifier, synonyms] (1189)
46. glucose intoleran*.mp. [mp=title, abstract, original title, name of substance word, subject heading word, floating sub-heading word, keyword heading word, organism supplementary concept word, protocol supplementary concept word, rare disease supplementary concept word, unique identifier, synonyms] (16545)
47. glucose toleran*.mp. [mp=title, abstract, original title, name of substance word, subject heading word, floating sub-heading word, keyword heading word, organism supplementary concept word, protocol supplementary concept word, rare disease supplementary concept word, unique identifier, synonyms] (59483)
48. obes*.mp. [mp=title, abstract, original title, name of substance word, subject heading word, floating sub-heading word, keyword heading word, organism supplementary concept word, protocol supplementary concept word, rare disease supplementary concept word, unique identifier, synonyms] (352263)
49. artheroscleros*.mp. [mp=title, abstract, original title, name of substance word, subject heading word, floating sub-heading word, keyword heading word, organism supplementary concept word, protocol supplementary concept word, rare disease supplementary concept word, unique identifier, synonyms] (149)
50. 28 or 29 or 30 or 31 or 32 or 33 or 34 or 35 or 36 or 37 or 38 or 39 or 40 or 41 or 42 or 43 or 44 or 45 or 46 or 47 or 48 or 49 (1781365)
51. 26 and 50 (337)
52. remove duplicates from 51 (334)
53. (infan* adj10 (body composition or body fat or fat percentage or fat % or lean mass or fat mass of fat free mass or muscle mass or grip strength or hand strength or antropometr* or skinfold or skinfold thickness or body mass index or BMI)).mp. [mp=title, abstract, original title, name of substance word, subject heading word, floating sub-heading word, keyword heading word, organism supplementary concept word, protocol supplementary concept word, rare disease supplementary concept word, unique identifier, synonyms] (3773)
54. ((baby or babies) adj10 (body composition or body fat or fat percentage or fat % or lean mass or fat mass of fat free mass or muscle mass or grip strength or hand strength or antropometr* or skinfold or skinfold thickness or body mass index or BMI)).mp. [mp=title, abstract, original title, name of substance word, subject heading word, floating sub-heading word, keyword heading word, organism supplementary concept word, protocol supplementary concept word, rare disease supplementary concept word, unique identifier, synonyms] (312)
55. (baby adj10 (body composition or body fat or fat percentage or fat % or lean mass or fat mass of fat free mass or muscle mass or grip strength or hand strength or antropometr* or skinfold or skinfold thickness or body mass index or BMI)).mp. [mp=title, abstract, original title, name of substance word, subject heading word, floating sub-heading word, keyword heading word, organism supplementary concept word, protocol supplementary concept word, rare disease supplementary concept word, unique identifier, synonyms] (131)
56. (babies adj10 (body composition or body fat or fat percentage or fat % or lean mass or fat mass of fat free mass or muscle mass or grip strength or hand strength or antropometr* or skinfold or skinfold thickness or body mass index or BMI)).mp. [mp=title, abstract, original title, name of substance word, subject heading word, floating sub-heading word, keyword heading word, organism supplementary concept word, protocol supplementary concept word, rare disease supplementary concept word, unique identifier, synonyms] (190)
57. ((child* preschool or preschool child) adj10 (body composition or body fat or fat percentage or fat % or lean mass or fat mass of fat free mass or muscle mass or grip strength or hand strength or antropometr* or skinfold or skinfold thickness or body mass index or BMI)).mp. [mp=title, abstract, original title, name of substance word, subject heading word, floating sub-heading word, keyword heading word, organism supplementary concept word, protocol supplementary concept word, rare disease supplementary concept word, unique identifier, synonyms] (670)
58. (early childhood adj10 (body composition or body fat or fat percentage or fat % or lean mass or fat mass of fat free mass or muscle mass or grip strength or hand strength or antropometr* or skinfold or skinfold thickness or body mass index or BMI)).mp. [mp=title, abstract, original title, name of substance word, subject heading word, floating sub-heading word, keyword heading word, organism supplementary concept word, protocol supplementary concept word, rare disease supplementary concept word, unique identifier, synonyms] (357)
59. (young children adj10 (body composition or body fat or fat percentage or fat % or lean mass or fat mass of fat free mass or muscle mass or grip strength or hand strength or antropometr* or skinfold or skinfold thickness or body mass index or BMI)).mp. [mp=title, abstract, original title, name of substance word, subject heading word, floating sub-heading word, keyword heading word, organism supplementary concept word, protocol supplementary concept word, rare disease supplementary concept word, unique identifier, synonyms] (269)
60. (kindergarten* adj10 (body composition or body fat or fat percentage or fat % or lean mass or fat mass of fat free mass or muscle mass or grip strength or hand strength or antropometr* or skinfold or skinfold thickness or body mass index or BMI)).mp. [mp=title, abstract, original title, name of substance word, subject heading word, floating sub-heading word, keyword heading word, organism supplementary concept word, protocol supplementary concept word, rare disease supplementary concept word, unique identifier, synonyms] (74)
61. (children under 5 adj10 (body composition or body fat or fat percentage or fat % or lean mass or fat mass of fat free mass or muscle mass or grip strength or hand strength or antropometr* or skinfold or skinfold thickness or body mass index or BMI)).mp. [mp=title, abstract, original title, name of substance word, subject heading word, floating sub-heading word, keyword heading word, organism supplementary concept word, protocol supplementary concept word, rare disease supplementary concept word, unique identifier, synonyms] (3)
62. (children under five adj10 (body composition or body fat or fat percentage or fat % or lean mass or fat mass of fat free mass or muscle mass or grip strength or hand strength or antropometr* or skinfold or skinfold thickness or body mass index or BMI)).mp. [mp=title, abstract, original title, name of substance word, subject heading word, floating sub-heading word, keyword heading word, organism supplementary concept word, protocol supplementary concept word, rare disease supplementary concept word, unique identifier, synonyms] (2)
63. (under 5’s adj10 (body composition or body fat or fat percentage or fat % or lean mass or fat mass of fat free mass or muscle mass or grip strength or hand strength or antropometr* or skinfold or skinfold thickness or body mass index or BMI)).mp. [mp=title, abstract, original title, name of substance word, subject heading word, floating sub-heading word, keyword heading word, organism supplementary concept word, protocol supplementary concept word, rare disease supplementary concept word, unique identifier, synonyms] (0)
64. 53 or 54 or 57 or 58 or 59 or 60 or 61 or 62 or 63 (5194)
65. 50 and 64 (2219)
66. (diabetes type 2 or type 2 diabetes or diabetes mellitus or insulin resistance syndrome* or insulin resistan* or hyperglycaemia or hypertension or arteriosclerosis or cardiovascular disease* or cardio vascular disease* or blood pressure or coronary heart disease* or metabolic syndrome* or dysmetabolic syndrome* or cardio metabolic disorder* or cardiometabolic disorder* or lipid profile or lipid metabolism or lipid* profile or glucose metaboli* disorder* or glucose intoleran* or glucose toleran* or obes* or artheroscleros*).mp. (1781365)
67. ((infan* or baby or babies or child* preschool or preschool child or early childhood or young children or kindergarten* or children under 5 or children under five or under 5’s) adj10 (body composition or body fat or fat percentage or fat % or lean mass or fat mass of fat free mass or muscle mass or grip strength or hand strength or anthropometr* or skinfold or skinfold thickness or body mass index or BMI)).mp. (5194)
68. 66 and 67 (2219)
69. ((infant* or baby or babies or child* preschool or preschool child or early childhood or young children or kindergarten* or children under 5 or children under five or under 5’s) adj10 (percent fat or fat percent or densitometry)).mp. [mp=title, abstract, original title, name of substance word, subject heading word, floating sub-heading word, keyword heading word, organism supplementary concept word, protocol supplementary concept word, rare disease supplementary concept word,unique identifier, synonyms] (71)
70. 67 or 69 (5242)
71. 66 and 70 (2221)
72. limit 71 to english language (2110)
73. limit 72 to humans (1911)
74. limit 73 to yr=“1990 -Current” (1817)
75. remove duplicates from 74 (1817)

## Appendix B

### Detailed summary of included studies

**Table B1.**
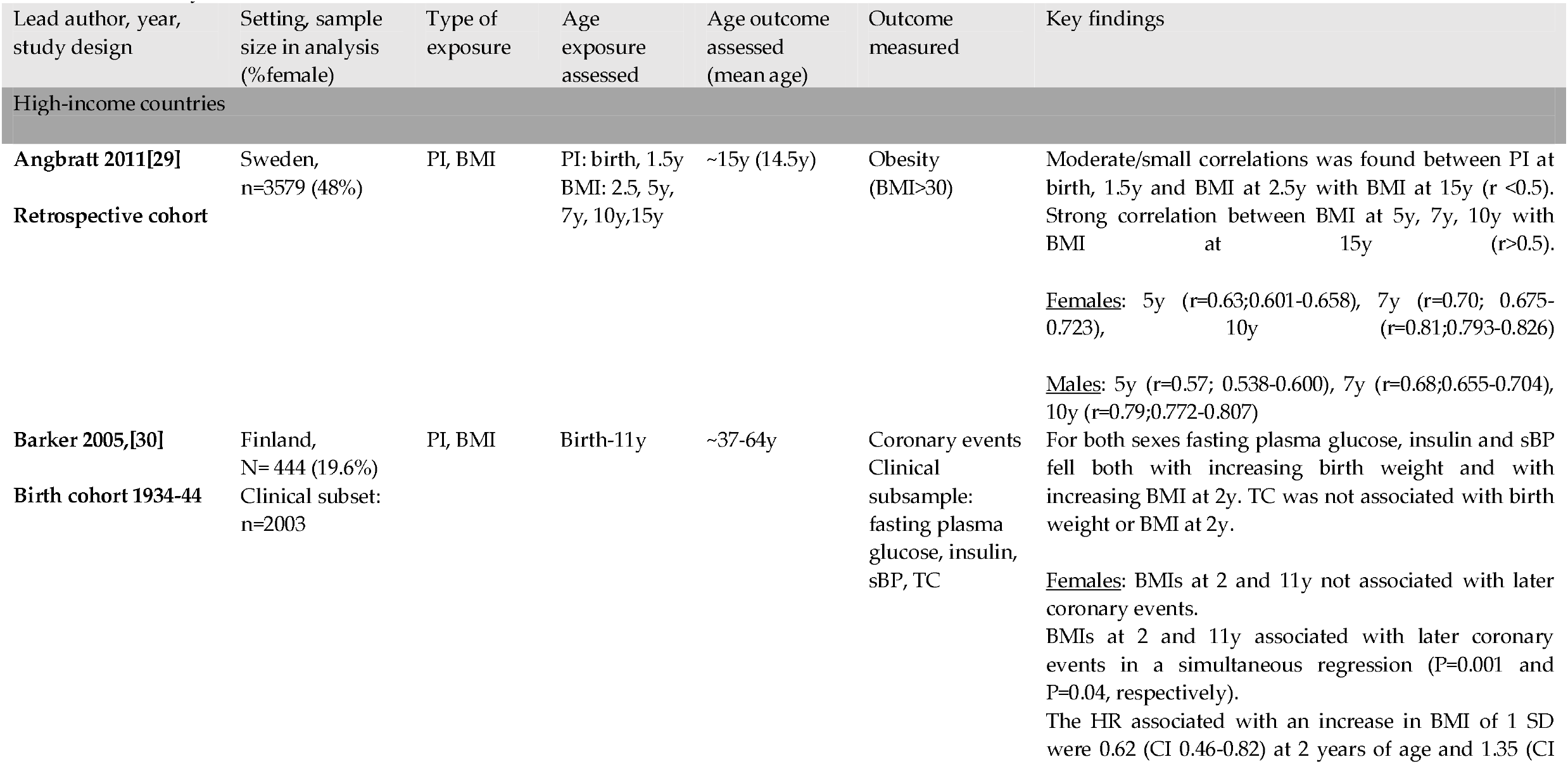
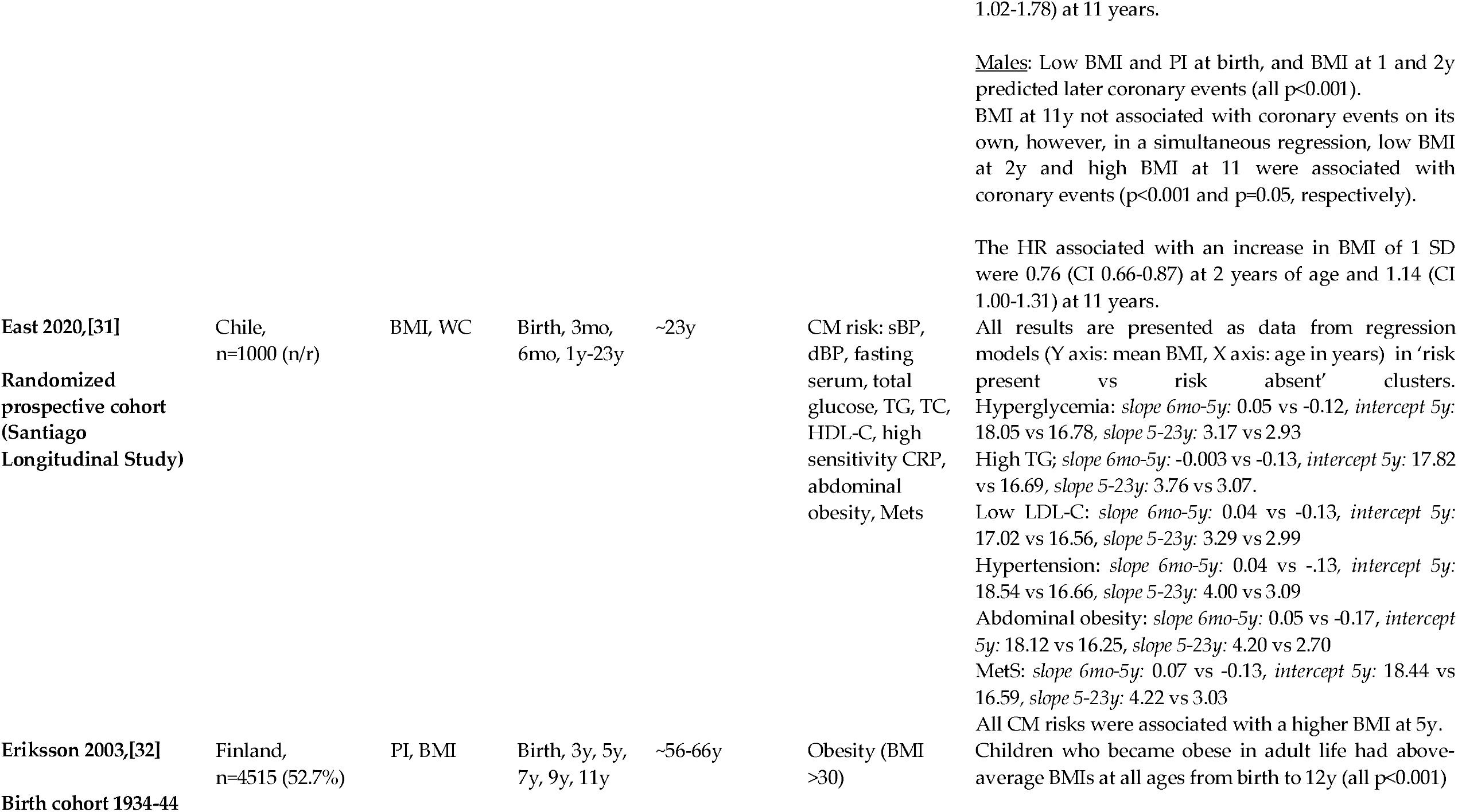
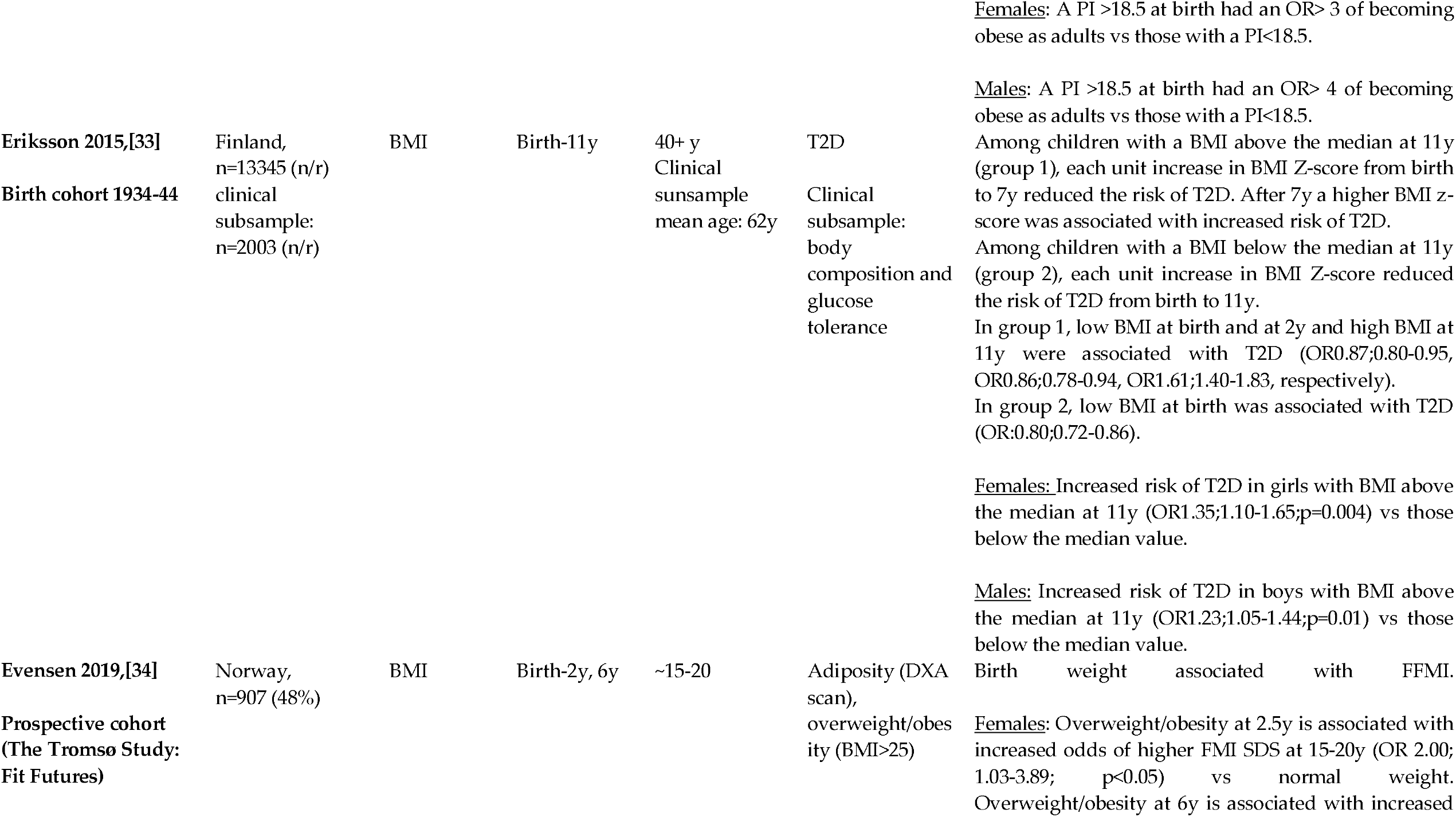
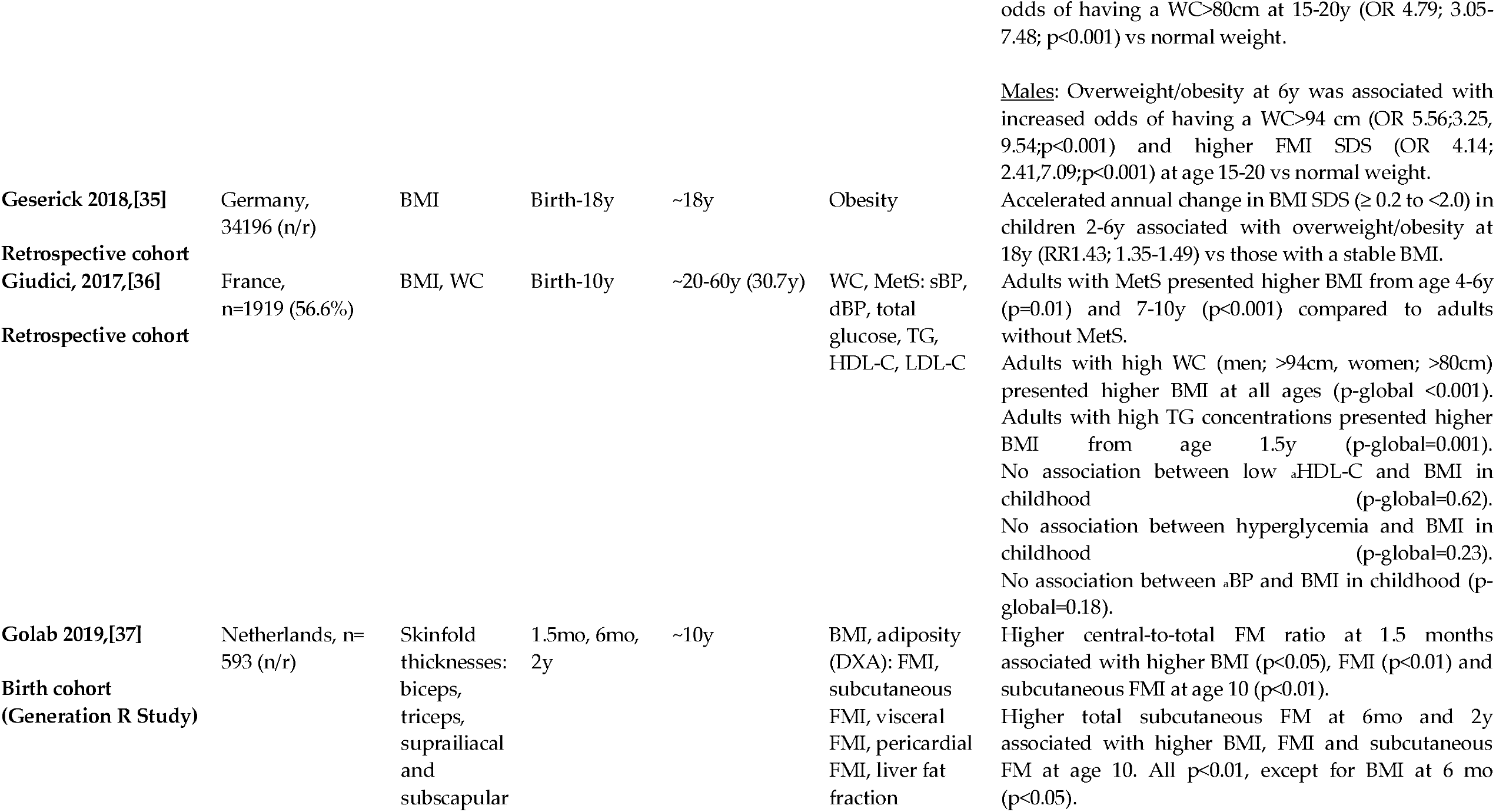
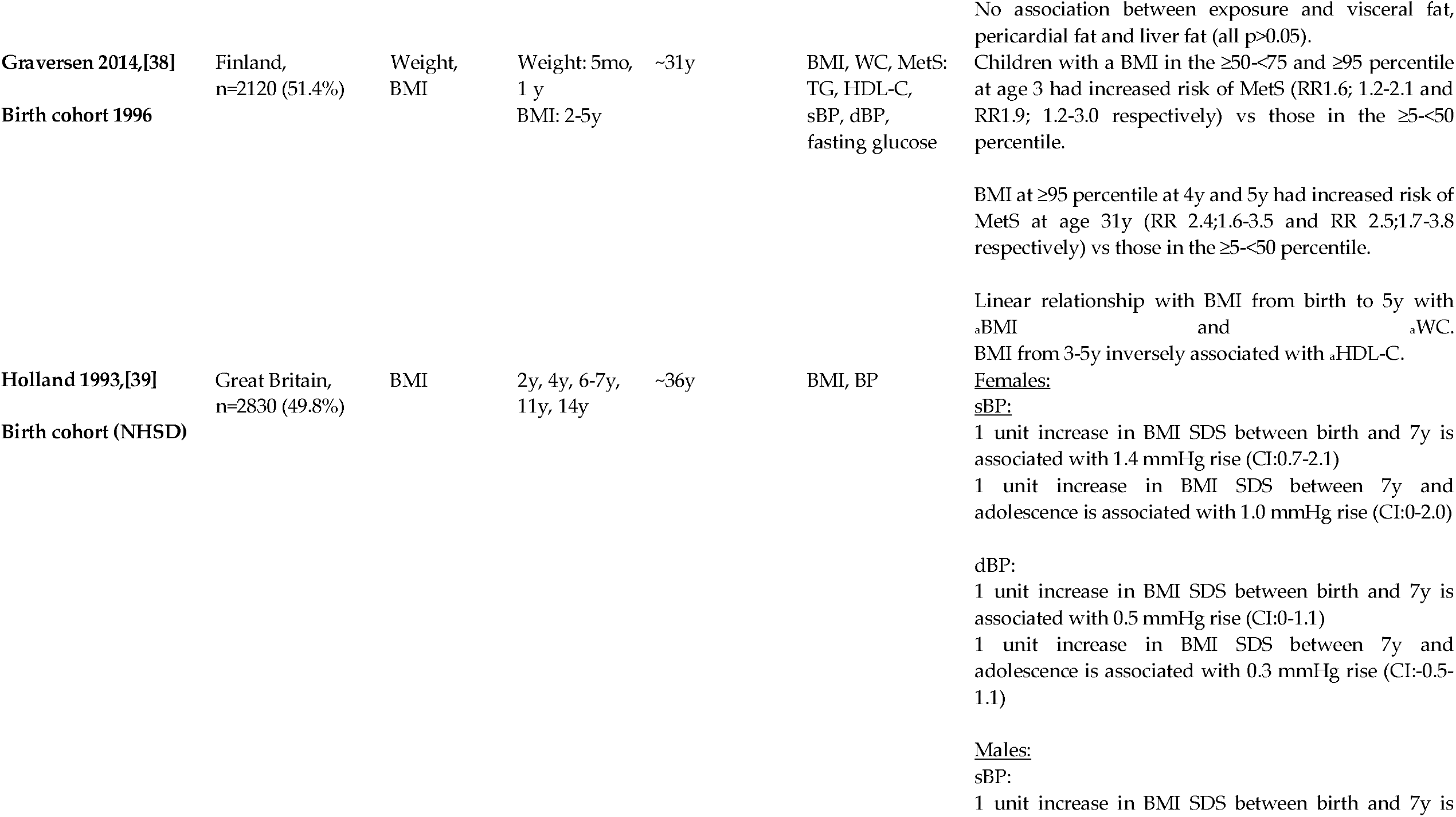
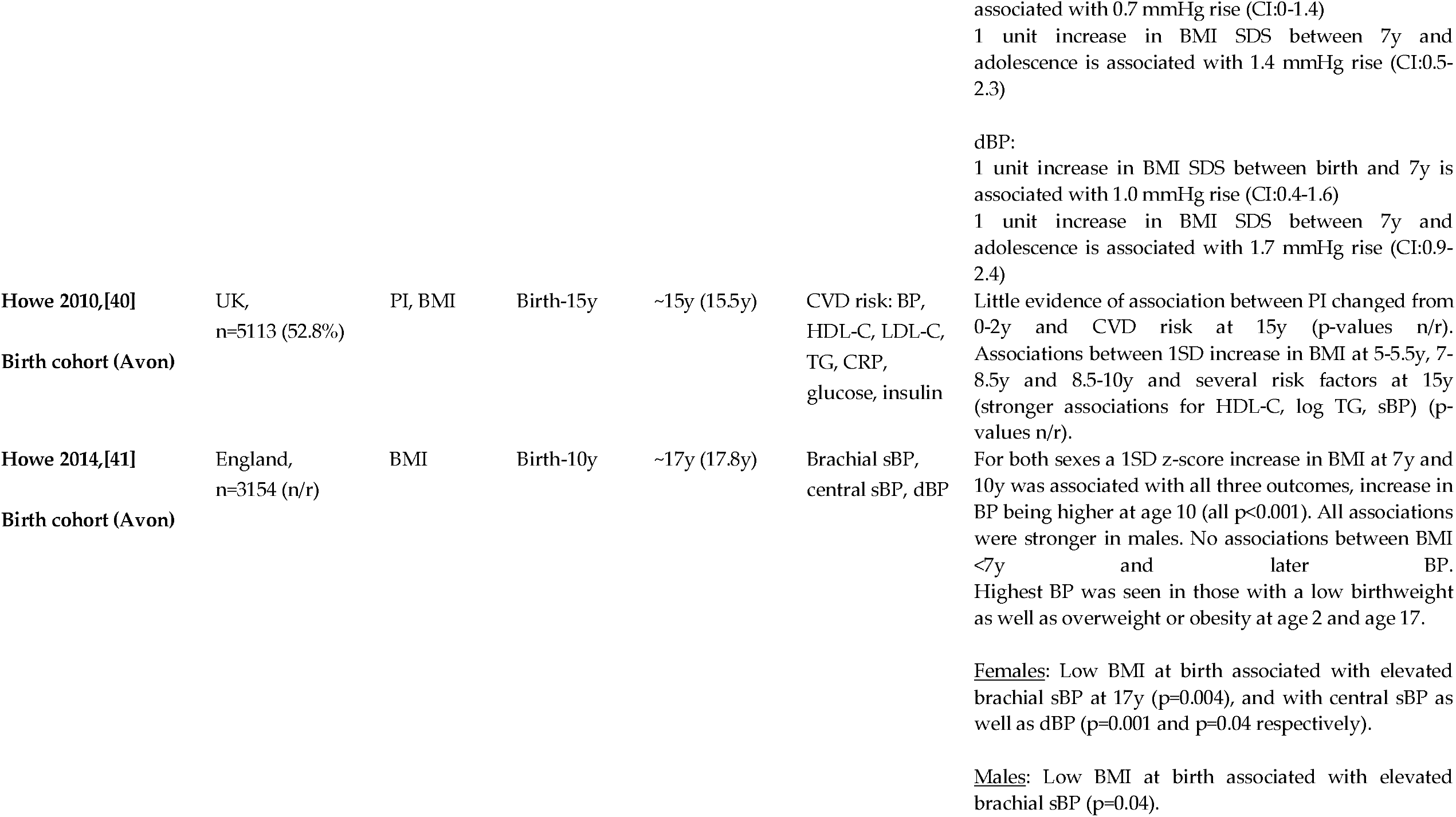
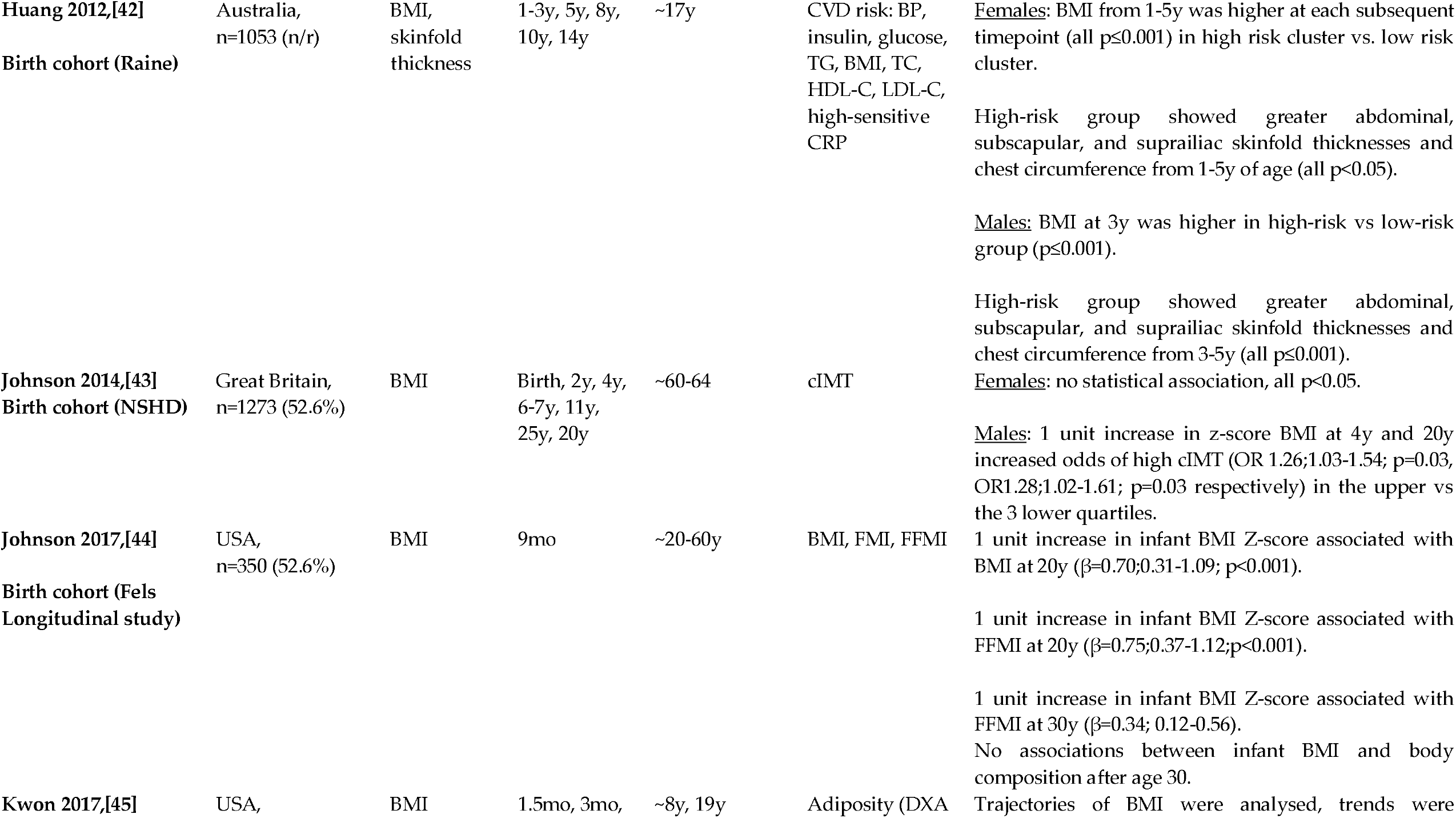
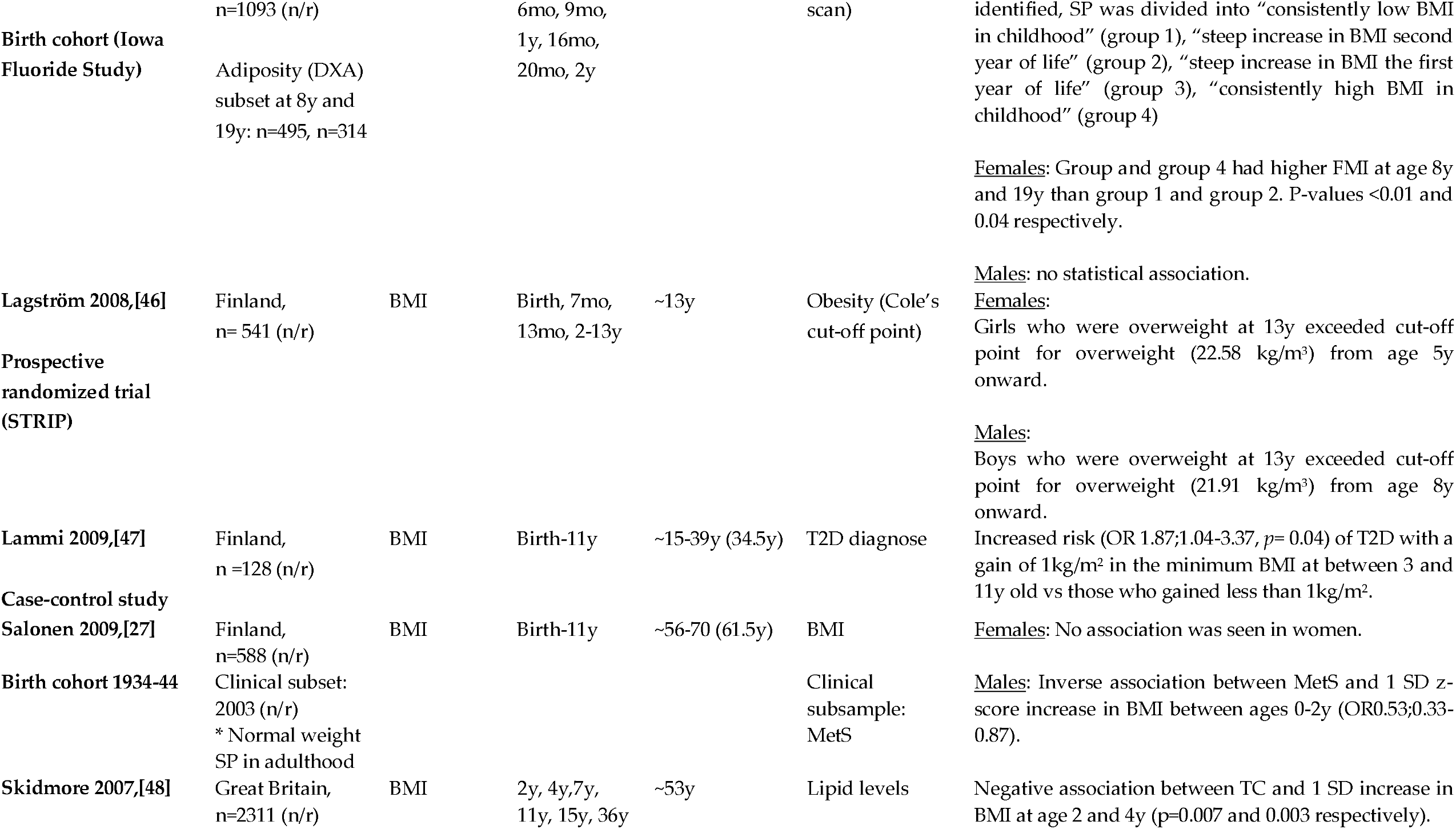
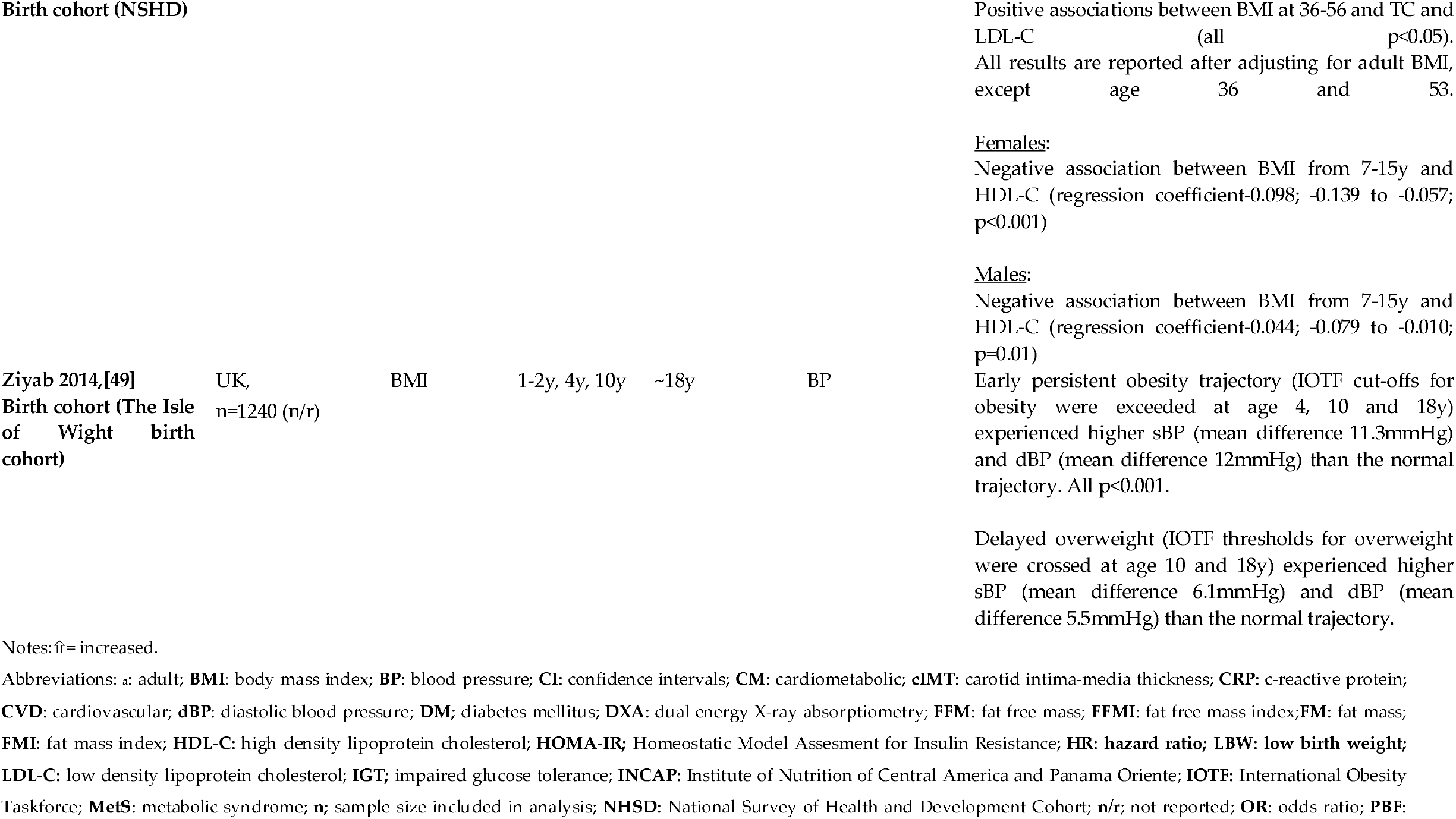
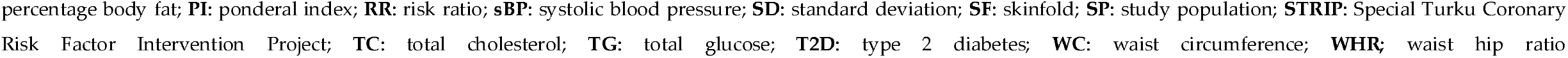
Full summary of included studies.

